# Effect of specific non-pharmaceutical intervention policies on SARS-CoV-2 transmission in the counties of the United States

**DOI:** 10.1101/2020.10.29.20221036

**Authors:** Bingyi Yang, Angkana T. Huang, Bernardo Garcia-Carreras, William E. Hart, Andrea Staid, Matt D.T. Hitchings, Elizabeth C. Lee, Chanelle J. Howe, Kyra H. Grantz, Amy Wesolowksi, Joseph Chadi Lemaitre, Susan Rattigan, Carlos Moreno, Brooke Borgert, Celeste Dale, Nicole Quigley, Andrew Cummings, Alizée McLorg, Kaelene LoMonaco, Sarah Schlossberg, Drew Barron-Kraus, Harrison Shrock, UFCOVID Interventions Team, Justin Lessler, Carl D. Laird, Derek A.T. Cummings

## Abstract

Non-pharmaceutical interventions (NPIs) remain the only widely available tool for controlling the ongoing SARS-CoV-2 pandemic. We estimated weekly values of the effective basic reproductive number (*R*_*eff*_) using a mechanistic metapopulation model and associated these with county-level characteristics and NPIs in the United States (US). Interventions that included school and leisure activities closure and nursing home visiting bans were all associated with an R_eff_ below 1 when combined with either stay at home orders (median *R*_*eff*_ 0.97, 95% confidence interval (CI) 0.58-1.39)* or face masks (median *R*_*eff*_ 0.97, 95% CI 0.58-1.39)*. While direct causal effects of interventions remain unclear, our results suggest that relaxation of some NPIs will need to be counterbalanced by continuation and/or implementation of others.

## Introduction

Months after the emergence of SARS-CoV-2 and its subsequent pandemic spread, widespread transmission continues. The United States (US) has been particularly affected, although the burden of disease has been geographically heterogeneous. While Northeastern states like New York, New Jersey, and Massachusetts were hardest hit when the virus first arrived in the US from March to April, 2020, states like Texas, Florida, Arizona, and California, which had previously avoided substantial disease burden, experienced rapidly growing cases between June and August (Fig. 1A) (*1*). Many factors likely contribute to spatial and temporal heterogeneity in COVID-19 incidence, including socio-demographic characteristics, the frequency of importation of infections, and the local use and timing of non-pharmaceutical interventions (NPIs) (*2*–*5*).

**Figure 1.**
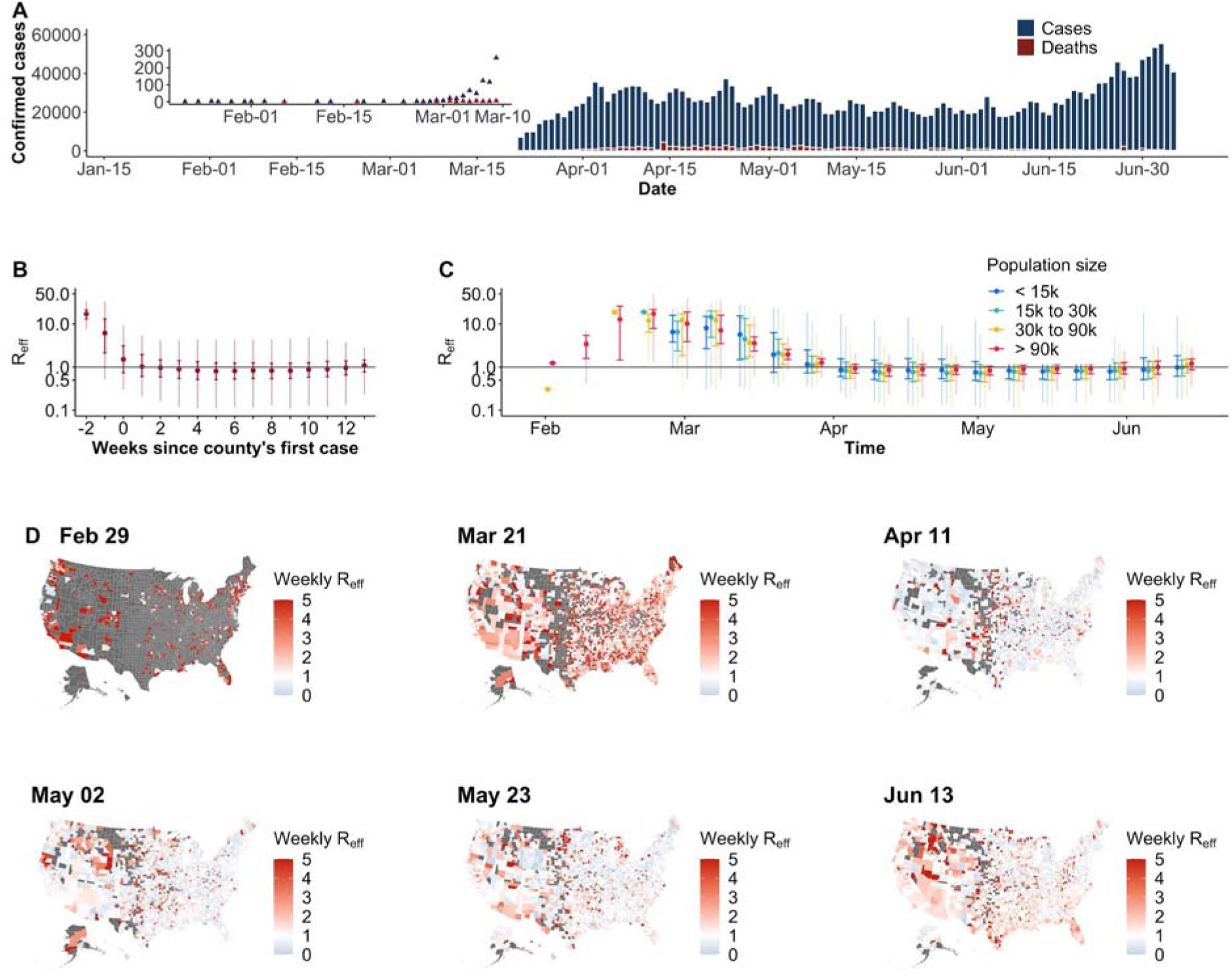
Confirmed COVID-19 cases and reproduction numbers (R_eff_) in the US. **A)** Daily confirmed COVID-19 cases and deaths. **B**) Distribution of R_eff_ by weeks since the county’s first reported case. Grey horizontal line indicates the threshold of R_eff_ = 1 (same as in C). Medians (points), interquartiles (dark vertical lines) and 95% percentiles (light vertical lines) are shown (same as in C). **C**) Temporal distribution of R_eff_ stratified by county population size. **D**) Map of county R_eff_ for representative weeks. Weeks were selected with a 3-week interval from the last week when R_eff_ were available. Grey indicates no data available.

While there is a continuing need to control SARS-CoV-2 transmission, prolonged social and economic strains push the limits of population compliance. Hence, it is critical to identify targeted and effective strategies for disease control. Variation in NPI application and timing across US counties provides an opportunity to compare the effectiveness of interventions in reducing transmission, but these effects are difficult to estimate directly due to spatial and temporal variation in transmission and an imperfect and ever-changing surveillance system. We hypothesized that by using multiple data streams and focusing on estimates of transmissibility rather than raw counts of cases or deaths, some of these challenges may be overcome.

Here, we estimated the transmissibility of SARS-CoV-2 in 3,036 (out of 3,142, 97%) US counties using a mechanistic meta-population model that incorporates spatial coupling of transmission between counties to estimate weekly effective basic reproductive numbers (i.e., R_eff_, the reproductive number adjusted for changes due to factors other than population susceptibility, such as social distancing) from confirmed cases and deaths from January 21 to July 5, 2020. We associated these R_eff_ estimates with NPIs and county level demographics, while accounting for, temporal variation, autocorrelation and uncertainty in our estimates. We aimed to determine which NPIs have been most effective so far to inform future implementations.

## Results

A total of 2,846,249 COVID-19 cases and 128,391 deaths were reported in the US as of July 5, 2020 (Fig. 1A). Cases first appeared in coastal counties with large populations in late-January and were reported in 68% of US counties by March 31. Weekly R_eff_ fit to confirmed cases (Fig. 1B-C) suggests that counties with larger population sizes (>90,000) experienced earlier and more efficient transmission (i.e. greater R_eff_) (median R_eff_ 2.6, interquartile range (IQR) 1.5 to 6.5 in the weeks before April) (Fig. 1C-D; Fig. S1). Later in the epidemic R_eff_ dropped in these large counties (median R_eff_ of 0.8, IQR 0.7 to 1.0 in the week ending May 2), and though R_eff_ was similar in small counties, some had appreciably higher transmission (median R_eff_ of 0.9, IQR 0.6 to 1.8) (Fig 1C-D).

We compiled detailed data on the timing of state level NPIs policies and county level data on school closure. Enactment of NPIs started in early March and peaked the week ending April 11, by which time 100% of counties had closed public schools; and, of states, 98% had closed leisure activities (restaurants, gyms and movie theaters), 88% had stay at home orders, 63% had suspended medical services, 59% had banned nursing home visits and 29% had closed daycares (Fig. 2A; Fig. S2). In most counties, the majority of interventions were implemented before a county had its first case; with school closure coming earliest (median 1.4 weeks before first case IQR 2.6 to 0.6 weeks) (Fig. 2B). The one exception is face masking orders which were initiated on average 5.7 weeks (IQR 4.1 to 7.0 weeks) after the first case (Fig. 2B). Many locations started to gradually lift control measures in mid-April, particularly medical service suspensions (remained in only one state as of June 13 (2%)), stay at home orders (12%) and leisure activities closures (45%) (Fig. 2A, S2). At time of analysis no county had lifted school closure.

**Figure 2.**
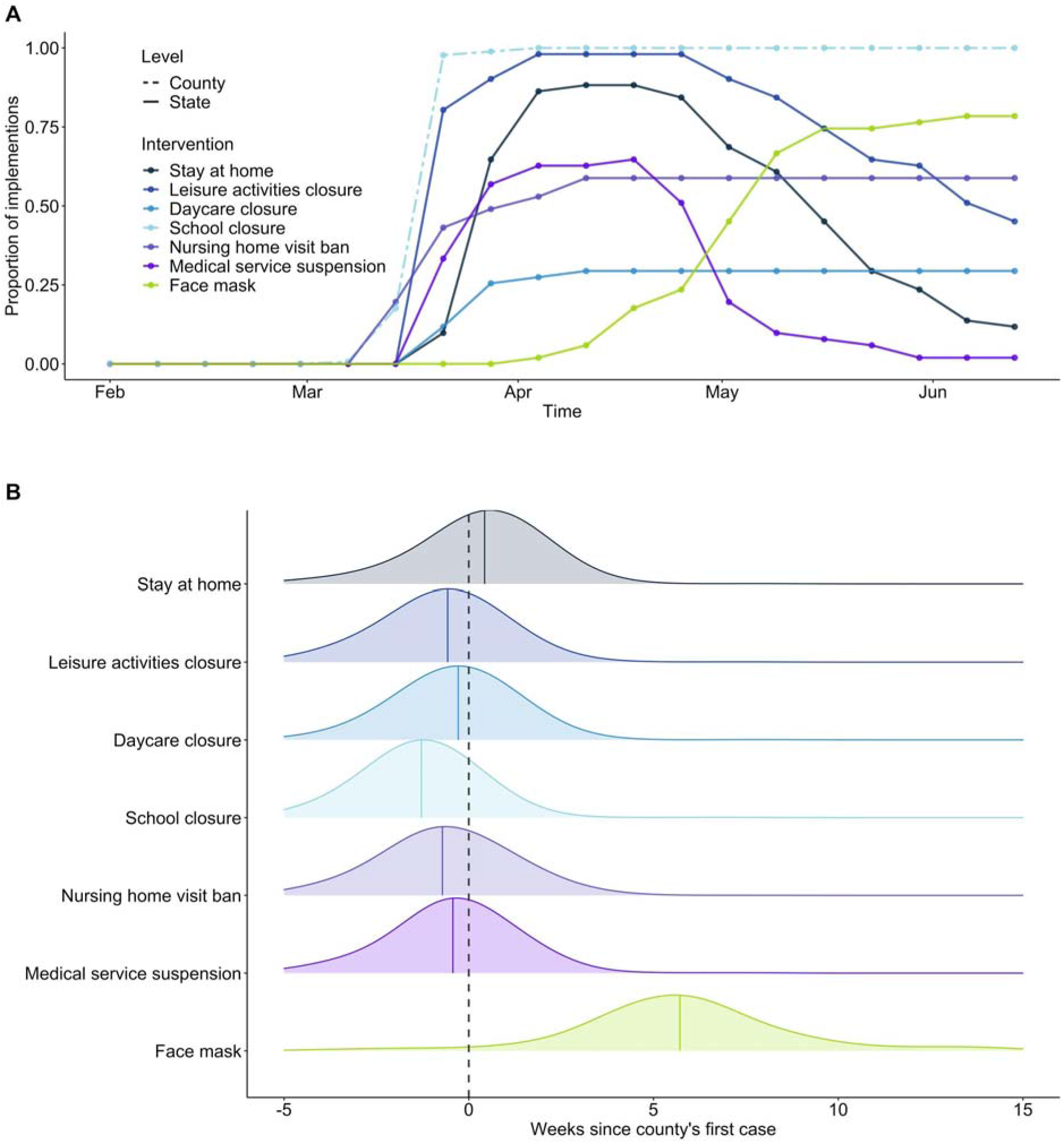
Temporal distributions of non-pharmaceutical interventions. **A**) Time series of the proportion of states and counties where interventions were implemented. **B)**Distribution of time differences between intervention times and the occurrence of the county’s first confirmed case. Colored, solid lines indicate the median difference times of each intervention.

In March, the average R_eff_ in each state was consistently high across the country (median 4.8, IQR 4.1 to 5.3), but transmission had reduced sharply by May (median 1.2, IQR 0.9 to 1.5). However, these results mask substantial variation within states at all points in the epidemic (mean intra-state coefficient of variation (CV) 1.39, IQR 1.28-1.44 in March, and 1.16, IQR 0.78-1.66 in May). Using generalized estimating equations (GEEs) to account for autocorrelation, we estimated the county-level effects of intervention policies on log-transformed R_eff_ adjusting for log-population size, proportion of individuals in poverty, median household income and other county level covariates (Tables S1-3; see Methods for description of all models considered). Transmission was consistently higher in counties with greater population sizes (21% increase in R_eff_ per 1,000 increase in population size, 95% CI 13 to 29%) and those with a higher proportion of people without college educations (5% per 10% increase, 95% CI 3 to 6%), while transmission was consistently lower in counties with higher proportions of white individuals (2.6% per 10% decrease in the proportion, 95% CI 2.0 to 3.2%) and those with a lower median age (0.6% per 1 year decrease in median age, 95% CI 0.3 to 0.9%).

We found that transmission had the strongest association with school closure (37% reduction in R_eff_, 95% CI 33 to 40%), followed by daycare closure (31%, 95% CI 26 to 35%) and banning nursing home visits (26%, 95% CI 23 to 29%) (Fig. 3A; main model in Tables S1-2). Stay at home orders were associated with a 15% reduction in R_eff_ (95% CI 13 to 17%) while face-mask orders were associated with a 18% reduction (95% CI 16% to 20%). Log-linear regression models that included a lag-term to account for autocorrelation (termed the lag model, an alternative approach to GEE) yielded similar regression coefficients (Fig. 3A and Table S4). To ensure the contribution of interventions in reducing R_eff_ when prioritizing parsimony, we fitted LASSO regression on the lag model model across a range of penalties for model complexity and found that school closure was the intervention most consistently associated with reductions in R_eff_ (Fig. 3B). Including information on NPIs improved model performance across multiple metrics (Tables S2, S3) (see Methods).

**Figure 3.**
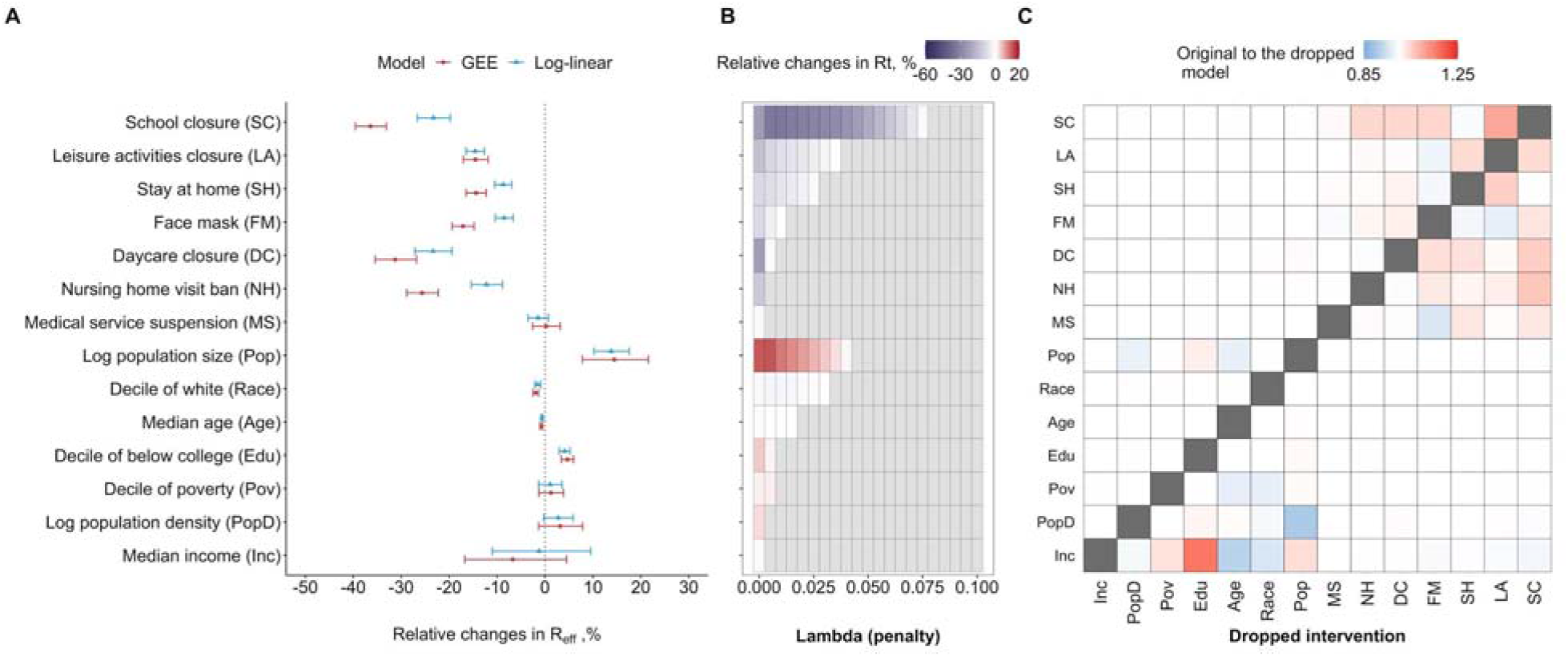
Effects of non-pharmaceutical interventions (NPIs) and county-level characteristics on transmission. **A**) Effects of NPIs and county-level characteristics estimated from the main model. Models were fitted with both GEEs and lag model regression. The order in y-axis (same for C) is according to the importance of covariates in explaining the variances shown in B. **B**) The importance of covariates in explaining the variances. Main models that were formulated for lag model regression were fitted to LASSO with increasing parsimony. **C**) Changes in the estimated effects when each covariate is dropped in the main model fitted with lag model regression.

Implementation of interventions was highly temporally correlated both between and within counties (Figs. 2, S2 and S3), presenting challenges to estimating independent associations. For example, school closures occurred at the same time in much of the country, with closure only reported in three unique weeks making separation of the effect of school closure from the effect of week of year impossible (including week of year as a categorical covariate eliminates the observed effect Fig. S4). However, whether causation, coincidence or confounding, reduction in R_eff_ occurred for some reason; hence, we performed several analyses to better disentangle the observed associations with NPIs.

Univariate and multivariate models for each NPI showed similar relative associations (Fig. S5) as did models that included an effect for time since the first case in a county (Fig. S6). To assess the impact of collinearities, we reran the main model holding out each NPI one at a time. The largest changes in estimated coefficients were seen when dropping school closure, which substantially impacted the estimated association of R_eff_ with remaining coefficients (e.g. the coefficient of banning nursing home visits indicated a 6.4% larger reduction in models without school closure) (Fig. 3C). To further ensure the observed associations between NPIs and reductions in R_eff_ were not merely the result of spurious associations between changes in R_eff_ and changes in NPI, we permuted data on NPIs across counties (Fig S7A,B) and within counties (Fig. S7C,D), respectively and found all associations to be substantially dampened or eliminated.

People changed their behavior in response to the pandemic, whether due to policy or personal choice. An important behavior that impacts SARS-CoV-2 transmission is travel to and social contact in different settings. Data on workplace presence relative to pre-pandemic periods were available on Google users (*6*) and, unlike other measures of movement in these data, were available for the nearly all (98%) of the county-weeks in our analysis. Across the US, large changes in workplace presentation occurred in March (Figure S8). We explored potential mediation/confounding by workplace presence in each county. We found that, at least in part, the relationship with a number of NPIs including school closure (32% [95% CI 28%, 34%] of total effect explained), leisure activities (65% [95% CI 59%, 71%] and stay at home orders (100% [95% CI 94%, 109%]) were mediated and/or confounded by workplace presence (Fig. S9, S10).

No single intervention was implemented alone for a sustained period of time in the period of our study, and many combinations of interventions never appear (e.g., there are no cases where stay at home orders are in effect but school closure is not). Hence, we only observe the impact of suites of interventions that may have complex interactions. We fit a GEE model to each of the unique suites of NPI utilized by counties (including no intervention) as categorical variables (Fig 4). We also fit a boosted regression tree model that can account for complex interactions between interventions (see Methods). Estimates from these models were consistent with those calculated from our main model. Interventions that included school and leisure activity closure and nursing home visiting bans were all associated with an R_eff_ below one when combined with either stay at home orders (median *R*_*eff*_ 0.97, 95% CI 0.58-1.39) or face masks (median *R*_*eff*_ 0.97, 95% CI 0.58-1.39) (Fig. 4). Inclusion of more interventions further reduced R_eff_, with a minimum median *R*_*eff*_ of 0.65 (0.39, 1.08) when all interventions are in place (Fig. 4, Table S7).

**Figure 4.**
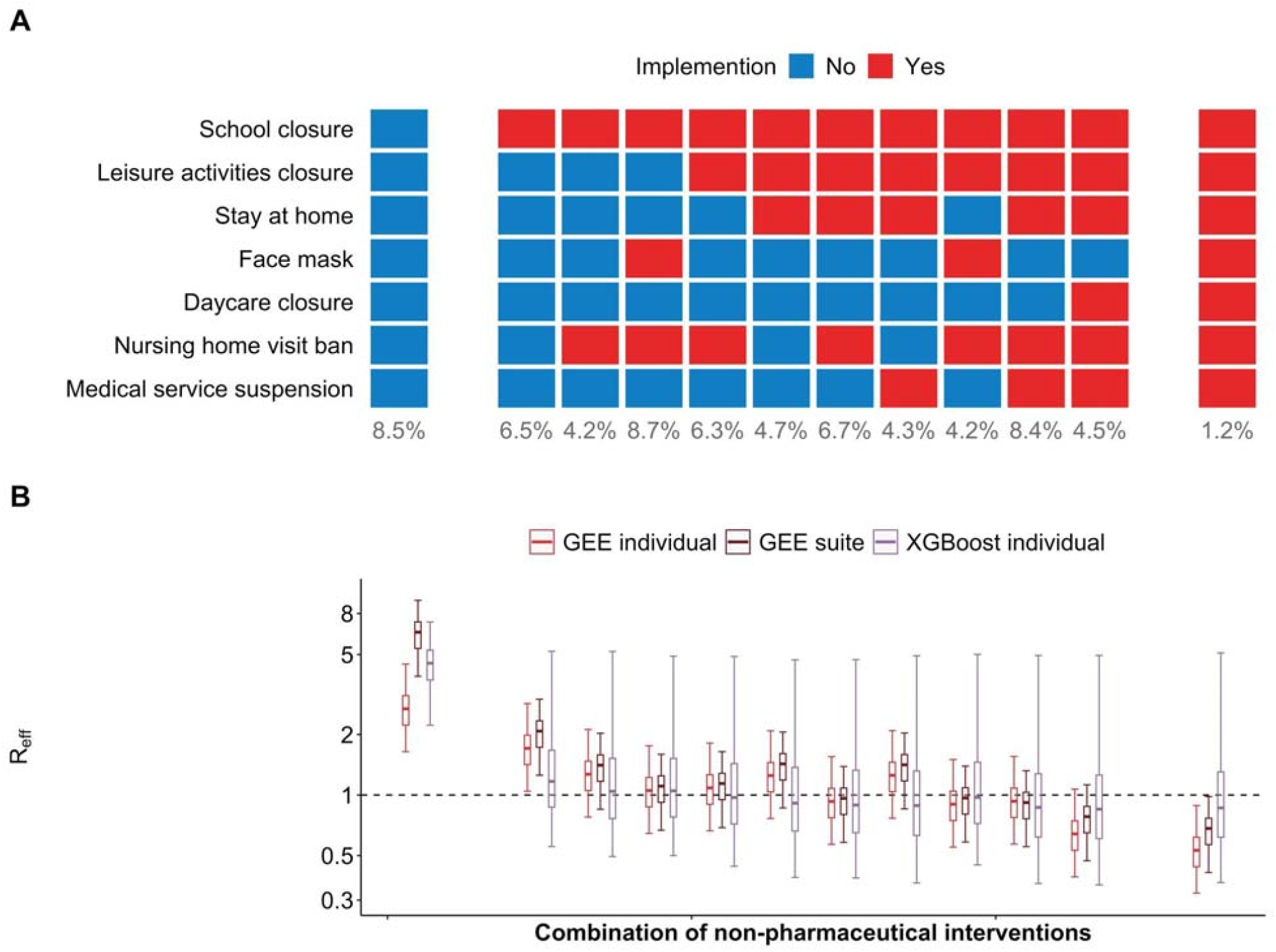
Prediction of reproduction numbers (R_eff_) associated with different non-pharmaceutical interventions (NPIs) suites. **A**) Non-pharmaceutical interventions (NPIs) suites: no intervention (left column), all interventions (right column) and the ten most frequently used NPI suites in counties in the US (middle columns; ordered by number of NPIs in each suite). Numbers below the columns are percentages of county-weeks in the study in which the suites were active. **B**) R_eff_ with different NPI suites. Median (cross), interquartile range (boxes) and 95% quantile range (vertical lines) of R_eff_ were distributions of predictions from the main model fitted with GEE (red, Fig. 3A), GEE with NPIs fitted as suites (brown), and boosted decision tree (purple). Horizontal dashed line indicates the threshold of one.

Multiple sensitivity analyses found broadly similar estimates of associations of NPI with R_eff_, these included: subsetting our analysis to exclude highly uncertain R_eff_ estimates from early in the pandemic (Fig. S11); allowing interventions to have an ongoing impact after they are lifted (Fig. S12); using alternative reconstruction methods (Fig. S13); and using estimates derived from reported deaths (Figs. S14). In the last analysis, we did observe reduced effects of most NPIs and a larger effect of school closure; though results were otherwise qualitatively the same.

## Discussion

A strength of our approach is the detailed data on NPIs that we compiled as well as the robust estimation of transmissibility using a mechanistic meta-population model. We found that six of seven NPIs examined were associated with reductions in R_eff_, with school closure being associated with the greatest reduction. However, we can only speculate about the mechanistic pathways by which any of these policies may have caused reductions in transmission. School closures, for example, may have had substantial impacts on the social interactions of non-school-aged individuals as parents and workplaces adapt to accommodate changes in children’s schedules, as suggested by our mobility analysis. It may also be a leading indicator of community attitudes about transmission. Our estimates indicate that four interventions were necessary, on average, to reduce R_eff_ below one, and that even with seven interventions, reproductive numbers remained above 0.65 on average (Fig 4). However, we note that our estimates do not include the effect of immunity due to substantial infection in many areas of the US, which will cause additional reductions in R. Overall, even when the greatest number of intervention policies were in place (only observed in 1.2% of county-weeks; Fig. 4A), we never saw reductions as large as those seen in Asia and parts of Europe, where reproductive rates fell as low as 0.44 (*7*). Whether this is the result of poor compliance, structural factors, or states easing restrictions before they could have their full effect remains unknown. However, even though reductions were not as large as those seen in other areas, stark and immediate reductions in reproduction numbers across the US coincided with the use of NPI.

We are not the first to attempt to estimate the impact of NPIs on transmission of SARS-CoV-2. Previous analyses agree with ours in several important dimensions, including the clear association of business closures, stay at home orders, and masking with significant reductions in transmission (*8, 9*). Beyond the impact of NPIs, other studies identified population density as a risk factor for transmission (*10*) while we found that county population size had a stronger association (Table S6).

There remains debate on the impact of school closure. Consistent with our results, Auger et al. (*2*) found significant associations between school closure and reduced incidence. However, others, e.g., Hsiang et al. 2020 (*8*), have found no association between school closure and the rate of growth in cases. These discrepancies may in part be explained by differences in how investigators treat the temporal association of interventions with changes in observed epidemic patterns (i.e., our estimates are lagged based on modeled epidemic processes and reporting delays), but the fact is that we never see an instance in the data where stay at home orders and many other effective interventions are implemented without school closure, so all other effects are measured in the presence of school closure. How this is handled in analysis can lead to substantial differences in estimated effect.

The impact of school closures need not necessarily stem directly from reductions in direct infection by school aged individuals. The associated reductions in R_eff_ we observed suggest substantial confounding or indirect effects of school closure. The association between school closure and time spent in the workplace provides one possible mechanism of indirect effects, however others exist. Further, even if transmission is rare schools bridge many communities, and can play an important role in facilitating epidemic spread by connecting these subpopulations. Regardless of why, the data and past experience show important associations between school closure and transmission which should not be dismissed when setting policy.

Aside from the correlation in timing of interventions, other factors may also challenge our inferences. Compliance with policies and lags between implementation and actions by individuals could obscure the impact of policies on transmission. School closure is unique in this regard among the NPI we considered as schools stayed closed with effectively 100% compliance over the period of our analysis. There may be confounders that were unmeasured or not included in our model. Though we were able to obtain county level data on school closures, we were limited to state level data on other interventions. Local changes that affected large populations within a state may lead to misclassification bias. Large spatial and temporal variation in the accuracy of surveillance for confirmed cases or deaths could induce spurious changes in R_eff_ that do not reflect true transmission (*11*), however using both cases and deaths for our inference helps mitigate this possibility. Many counties reported limited numbers of cases and/or deaths and thus infection dynamics could not be reconstructed. We assumed a stable distribution of delays between infection and the time of confirmation or death, though this could have varied over the course of the outbreak (*12*). Not all behavioral change is captured by concrete policies; however, our focus was on the impact of policies enacted by governments rather than actions taken by individuals in the absence of such policies.

Despite these limitations our analysis provides critical insight into how individual interventions, or at least commonly used suites of interventions, may affect the spread of SARS-CoV2. These estimates are critical as governments attempt to figure out how to respond to resurgent cases and look for responses that successfully control spread while allowing as much of normal economic and social life to continue as possible. The fact that multiple NPIs were needed to reduce R_eff_ below one, suggests that relaxation of some NPIs might need to be counterbalanced by continuation and/or implementation of others. Our point estimates of the relative contribution of each intervention provide some guidance in making these difficult decisions.

## Materials and Methods

Data and code for analysis are made available at https://github.com/datcummings/SARS-CoV-2.NPI.analysis.

### Data on COVID-19 cases and deaths

Laboratory-confirmed COVID-19 cases and deaths in the counties of the United States were retrieved from USAFacts (https://usafacts.org/visualizations/coronavirus-covid-19-spread-map/) on July 5, 2020. The data from USA Facts was used for all counties except for New Mexico, where we observed timing offsets in weekend reporting for a few, select dates and counties. Data from the NY Times did not have these issues and thus were used for New Mexico (1).

### Data on county-level demographic characters

We obtained data on county-level demographic characteristics (i.e. population size, population median age, land area, median annual income, number in poverty, number reported as white in race, and number with education below college) from the 2014-2018 American Community Survey, which were retrieved through *tidycensus* package version 0.9.9.2 (*13*) in R. Population density was calculated by dividing the population size over land area. Proportions of population in poverty, white in race, and education below college were derived by dividing the numbers with the total number of people surveyed for those characteristics.

#### Data on non-pharmaceutical interventions (NPIs)

We obtained dates of policy announcements of closures of public schools (K-12) of each county by consulting the government websites of school district, county and state and local news sources. We used the earliest documented date as the county’s school closure date when there were multiple dates available for districts within that county. When closures were announced during or at the end of a planned school break, the date when schools were last in session was reported. We found school closure dates for 94.3% (2,963 out of 3,142) of counties. We limited our analysis to the 2,963 counties for which school closure information was available (from the 3,036 for which we had estimates of R_t_).

We obtained dates of implementation and termination of the other NPIs at state-level from COVID-19 US State Policy Database (CUSP) on July 6, 2020 (*14*). Eleven interventions that were directly used to reduce transmissions were extracted from the dataset (Table S5). Interventions were grouped if they were semantically similar and clustered in time (e.g. face mask mandated in the public and face mask mandated in businesses; Figure S11). When multiple dates of interventions and terminations were available for the grouped NPIs, we used the earliest documented date for implementations and the latest date for termination. An intervention is considered as implemented if the date of the corresponding R_eff_ was between its implementation and termination date.

### Estimating reproduction number

We fit a mechanistic transmission model to confirmed cases of COVID-19 in each of the counties of the US. Separately, we also fit these same models to deaths due to COVID-19 in each county. We used spatially coupled SEIR models to represent the transmission of SARS-CoV-2 with separate metapopulations representing the population in each US county. We estimated the incidence of infection in each county based on confirmed count data (and separately deaths due to COVID-19) using the back-projection method of Becker et al (*15*). For our deconvolution procedure, numbers of cases and deaths were upscaled by sampling from a negative binomial (with probability 1/10 and 1/200 respectively). Second, using the delay distributions for cases and deaths, we applied the function ‘backprojNP’ from package ‘surveillance’ in R to back-project incidence. Finally, to account for right-truncation of incidence values, we followed Abbott et al (*16*) and sampled the estimated incidence to account for infections that had occurred but had not yet been reported or confirmed from a negative binomial distribution, where the probability was given by the respective cumulative delay distributions. The most recent 14 days were then removed. We constructed 100 stochastic realizations of this algorithm for each county. Using the resultant time series of daily infections in each county, we fit our mechanistic model with state variables S (susceptible to SARS-CoV-2), E (exposed, infected but not yet infectious), I (infectious), and R (recovered or deceased and no longer infectious) to each stochastic realization using non-linear optimization tools (IPOPT, Pyomo)(*17, 18*). Models were developed using epi_inference, a software package to be released. Our model included three infectious components to allow the distribution of durations of infectiousness to approximate a gamma distribution. Fitted parameters were piecewise constant transmission coefficients in each county for each week. Estimates used two weeks of incident infections to derive each piecewise constant transmission coefficient in each week (with estimates assigned to the first week of the two weeks used to estimate infections). A mobility matrix derived from US Census commuting data from pre-pandemic time periods was used to specify the fraction of commuters in each county and the fraction of time that those commuters spent in counties other than the ones they reside.

### Natural History of SARS-CoV-2

Time delays from infection to confirmation among those cases that are confirmed was assumed to be log-normally distributed with mean of days (log(8)=2.07 days). This assumption was derived from (*19*) and assumed confirmation comes on average 1.5 days after attendance at a medical facility. We assumed a log-standard deviation of 0.3 of this distribution (*20*). Time delays from infection to death among those that die was assumed to be log-normally distributed with log-mean 2.84 and log-standard deviation of 0.72 and assuming no competing risks (*21, 22*).

### Estimating effects of NPIs on transmission

In general, models that included different sets of covariates (i.e. autoregression of R_eff_, additional temporal marker for county-time, county-level demographic characteristics and NPIs; see details in Table S1 and S2) were fitted with generalized estimating equations (GEEs; *geeglm* of package “geepack”) and autoregressive lag model regressions, separately. We included AR(1) in GEEs and included log R_eff_ in previous week in lag model regression to account for autoregression of R_eff_. We weighted log-R_eff_ with its inverse coefficient of variation across the above-mentioned 100 stochastic reconstructions. NPI were included as a time-varying covariates with the status of the NPI defined for each week of the analysis as either 1 if in use in that county in a particular week or 0 if not. Table S1 describes the components of the **main** model and alternatives, **base**, the model with the minimum number of covariates that we considered, **time**, which included the covariates included in **base**, but adding time since the first case in each county as a categorical variable and **time and interventions**, which adds NPI to the **time** model.

In our main results, we interpreted the estimated effects of NPIs from the main model that was fitted with GEEs (Fig. 3A). Autoregression of R_eff_, county-level characteristics and NPIs were included in the main model (Table S1). To examine the reduction of different combinations of NPIs on R_eff_ (Fig. 4), we calculated R_eff_ by combining effects for individual NPIs that were estimated from the main model. To further examine whether the effects of NPI suites were robust to interactions between NPIs, we 1) fitted another model with GEEs by including autoregression of R_eff_, county-level characteristics and NPIs suites (as categorical variables); and 2) fitted a model with XGBoost of individual NPIs (details described below). We then compared the predictions on the R_eff_ for NPIs suites from the GEE suite model and XGBoost model to those calculated from our main model (Fig. 4B).

### Sensitivity analyses of effects of NPIs on transmission

#### LASSO

To examine the relative importance of NPIs in explaining variance of R_eff_ with increasing parsimony of the model, we performed LASSO (*glmnet* in “glmnet” package) with the main model that was fitted with our lag model regression. The estimated effects of NPIs from lag model regression are highly correlated with those estimated from GEEs (Fig. 3A). Effect sizes of NPIs were presented as the complexity penalizing hyperparameter (A) was increased from 0.0 to 0.1 at intervals of 0.005 (Fig. 3B). Coefficients estimated when A is 0 (i.e. no penalty) are equivalent to the estimates from lag model regressions shown in Fig. 3A.

#### Collinearity of covariates

We assessed the potential impact of colinearity of NPIs on our estimated associations of NPI with R_eff_ through two approaches: dropping one NPI at a time (“dropped model” hereafter) prior to fitting the model, and by fitting a single-intervention model. For the dropped model, we dropped individual covariates of NPIs and county characteristics one at a time and reran the main model that was fitted to lag model regression. We calculated the ratio of relative changes in R_eff_ that was estimated from the main analysis to that estimated from the dropped models (Fig. 3C). For the single-intervention model, we added one NPI into the base model at a time and compared the estimated effect of that NPI with the original effect size that was estimated from the main model. Single-intervention models were also fitted to GEEs and lag model regression (Fig. S5).

#### Impact of temporal declining trend of R_*eff*_ *on estimating effect of NPIs*

In competing NPIs against temporal markers, we further included a fixed-effect associated with county-time (i.e. time since a county had its first case) into the main model as a weekly discretized coefficient that was shared across all counties (Table S1 and S2). Weeks since the first case in a county were computed for each data point and were included in the models as categorical variables, i.e. from three weeks before to thirteen weeks after the county saw its first case.

In order to examine whether the effect of NPIs estimated from our main models was not just capturing the declining trends of R_eff_ over time, we permuted NPIs spatially (i.e. permuted the NPIs suites between counties) and temporally (i.e. permuted the time series of NPIs within counties) and refit the main model. We performed the permutations tests 100 times.

#### Impact of uncertainty from early transmission phase

R_eff_ estimates at the beginning of an outbreak are often challenged by large uncertainties. In order to examine the effects of these uncertainties on our estimated effects of NPIs, we fitted the main model to a subset of our data set, which only includes R_eff_ since two weeks after the county saw its first case.

### Effects of NPIs relaxations on transmission

In order to look at the effects of relaxing NPIs on the transmission, we fitted the main model by further splitting the effect of NPIs into “during the implementation” (intervention on) and “after the implementation was lifted” (intervention off). Relaxations were available for non-essential business, stay at home and medical service suspension. The rest of the covariates were the same as in the main model.

### Using deaths as alternative data to estimate R_eff_

R_eff_ that were estimated from the deconvoluted COVID-19 deaths were derived to assess the robustness of our statistical inference to using these data compared to confirmed cases. We therefore used the above-mentioned methods to estimate weekly R_eff_ from confirmed deaths and refit the main models with GEEs and lag model regression (Fig. S9). R_eff_ were estimated for 1,840 out of 3,142 counties (58.6%).

### Out-of-sample prediction

Models were fitted with each of the fifty states and District of Columbia held out as test sets. Prediction performances were measured using root mean squared error (RMSE), mean absolute scaled error (MASE), and coefficient of determination (R^2^). Fitting procedures for the lag models were as described above.

#### Comparative model

We employed XGBoost (*23*), a decision tree boosting package, to explore whether more predictive power can be gained through complex model structures. Optimal values of three main hyperparameters (fraction of covariates included in each boosting iteration, fraction of training data included per those iterations, and maximum tree depth) were determined through grid search; ranges (and grid intervals) were 0.3 to 0.9 (0.1), 0.3 to 0.9 (0.1), and 3 to 9 (1), respectively. Performances were evaluated under 10-fold cross-validation. Learning rate was conservatively set to 0.2 and the maximum number of iterations was capped at 200 with early stopping if RMSE does not improve after two iterations to avoid overfitting. The respective optimal values were 0.9, 0.3, and 6. We further optimized the maximum iterations cap when test sets were held out by spatial units; range of 50 to 200 at intervals of 25. Results reported were from the optimal iteration of 75 (which minimized RMSE).

### Effects of NPIs mediated by workplace attendance

Google Community Mobility Reports (https://www.google.com/covid19/mobility) were downloaded on September 16, 2020. Mean of daily percentage change of work commutes relative to the baseline of each county were computed for each weekly interval where we have R_eff_ estimates. We chose to focus on workplace presentation among candidate datasets available through Google as this was the only dataset that had less than 50% of county-weeks missing. Mediation analyses were conducted for each NPI separately. For each NPI, we fitted a full model to log transformed weekly R_eff_ and adjusted for county-level characteristics, the workplace attendance and the examined NPI. We then fitted a mediation model which regresses the workplace attendance on the examined NPI. The mediation analyses were conducted using R package “*mediation*” (*24*).

## Disclaimers

Sandia National Laboratories is a multimission laboratory managed and operated by National Technology and Engineering Solutions of Sandia LLC, a wholly owned subsidiary of Honeywell International Inc. for the U.S. Department of Energy’s National Nuclear Security Administration under contract DE-NA0003525. This paper describes objective technical results and analysis. Any subjective views or opinions that might be expressed in the paper do not necessarily represent the views of the USDOE or the United States Government. Contributions from Sandia National Laboratories were partially funded by the Laboratory Directed Research and Development (LDRD) program, and the DOE Office of Science through the National Virtual Biotechnology Laboratory, a consortium of DOE national laboratories focused on response to COVID-19, with funding provided by the Coronavirus CARES Act.

Material has been reviewed by the Walter Reed Army Institute of Research. There is no objection to its presentation and/or publication. The opinions or assertions contained herein are the private views of the author (AH), and are not to be construed as official, or as reflecting true views of the Department of the Army or the Department of Defense.

