## Supplemental Material for "Effect of specific non-pharmaceutical intervention policies on SARS-CoV-2 transmission in the counties of the United States"

### Supplementary Tables

**Table S1.** Covariates included in the main model and sensitivity analyses.

| **Covariates** | | | **Whether or not included in the model** | | | |
| --- | --- | --- | --- | --- | --- | --- |
| **Category** | **Detailed covariate** | | **Base** | **Main** | **Time only** | **Time and interventions** |
| **Autoregressive** | | AR(1) for GEEs;  Log_10_ Reff_-1_ for lag model | Yes | Yes | Yes | Yes |
| **County time** | Week -3 to 13 (categorical) | | No | No | Yes | Yes |
| **County characteristics** | State-specific intercept | | Yes | Yes | Yes | Yes |
|  | Median income | |  |  |  |  |
|  | Median age | |  |  |  |  |
|  | Log_10_ population size | |  |  |  |  |
|  | Log_10_ population density | |  |  |  |  |
|  | Decile of poverty | |  |  |  |  |
|  | Decile of white | |  |  |  |  |
|  | Decile of no college education | |  |  |  |  |
| **NPIs** | School closure | | No | Yes | No | Yes |
|  | Leisure activities closure | |  |  |  |  |
|  | Stay at home | |  |  |  |  |
|  | Daycare closure | |  |  |  |  |
|  | Nursing home visit ban | |  |  |  |  |
|  | Medical service suspension | |  |  |  |  |
|  | Face mask | |  |  |  |  |

**Tables S2.** Comparison of models with temporal markers and non-pharmaceutical interventions.

| **Model** | **Covariates included^#^** | | **GEE**^†^ | | **Lag Model**^§^ | |
| --- | --- | --- | --- | --- | --- | --- |
|  | **Additional temporal marker** | **NPIs** | **Adj. R^2*^** | **ΔQIC** | **Adj. R^2^** | **ΔAIC** |
| **Base** | -- | No | 7.5% | Ref | 25.0% | Ref |
| **Main** | -- | Yes | 21.8% | -336.3 | 29.0% | -1681.9 |
| **Time only** | Weeks since county’s first case (categorical) | No | 18.8% | -258.8 | 26.3% | -537.0 |
| **Time and interventions** | Weeks since county’s first case (categorical) | Yes | 24.1% | -329.8 | 29.5% | -1908.2 |

^#^ All models include county-level census information (i.e. population size, median income and decimal of poverty).

^†^ Autocorrelation of Reff was adjusted using a correlation structure of AR(1) in the models.

^§^ Autocorrelation of Reff was adjusted by including prior week’s Reff as a covariate in the models.

**Tables S3.** Prediction performance evaluated through out-of-sample validation compared against a comparator high-complexity model given equivalent sets of covariates.

| **Model type** | **Median performance (95% IQR)** | | |
| --- | --- | --- | --- |
|  | **RMSE** | **MASE** | **R^2^** |
| Main model  (Lag model) | 0.31  (0.27, 0.36) | 0.83  (0.81, 0.86) | 0.21  (0.15, 0.29) |
| Comparator model (boosted decision trees) | 0.33 (0.13, 0.44) | 0.84 (0.75, 0.97) | 0.22 (0.03, 0.75) |

* One of the fifty US states or District of Columbia was held out from the training set each time. Performances were computed from the held out data, i.e. the test set. Metrics measured are root mean squared error (RMSE), mean absolute scaled error (MASE) and coefficient of determination (R^2^). Values are given as the median across the 51 spatial units with 95% interquartiles in parentheses.

**Table S4.** Effects of non-pharmaceutical interventions (NPIs) and county-level characteristics on transmission. Estimates of the proportional reduction associated with each NPI or county-level characteristic for both GEE and lag model regressions.

| **Variable** | **GEEs** | **Lag model** |
| --- | --- | --- |
| School closure | 0.37 (0.33, 0.40) | 0.23 (0.20, 0.27) |
| Leisure activities closure | 0.14 (0.11, 0.16) | 0.14 (0.12, 0.16) |
| Stay at home | 0.15 (0.13, 0.17) | 0.09 (0.07, 0.11) |
| Face mask | 0.18 (0.16, 0.20) | 0.09 (0.07, 0.11) |
| Daycare closure | 0.31 (0.26, 0.35) | 0.23 (0.19, 0.27) |
| Nursing home visit ban | 0.26 (0.23, 0.29) | 0.12 (0.09, 0.16) |
| Medical service suspension | 0.00 (-0.03, 0.03) | 0.02 (-0.00, 0.04) |
| Log population size | -0.16 (-0.24, -0.09) | -0.14 (-0.18, -0.11) |
| Decile of white | 0.02 (0.01, 0.02) | 0.01 (0.01, 0.02) |
| Median age | 0.01 (0.01, 0.01) | 0.01 (0.00, 0.01) |
| Decile of below college | -0.05 (-0.06, -0.04) | -0.04 (-0.05, -0.03) |
| Decile of poverty | -0.01 (-0.04, 0.02) | -0.01 (-0.03, 0.02) |
| Log population density | -0.03 (-0.08, 0.02) | -0.02 (-0.05, 0.01) |
| Median income | 0.07 (-0.05, 0.17) | 0.02 (-0.09, 0.12) |

**Table S5.** Groupings of non-pharmaceutical interventions (NPIs) extracted from the COVID-19 US State Policy Database (CUSP)(*14*) on July 6, 2020.

| **Interventions in dataset ^#^** | **Groupings of interventions^†^** |
| --- | --- |
| School closure | School closure |
| Daycare closure | Daycare closure |
| Nursing home visit ban | Nursing home visit ban |
| Face mask mandated in public | Face mask |
| Face mask mandated in businesses |  |
| Close restaurants | Leisure activities closure |
| Close gyms |  |
| Close movie theaters |  |
| Stay at home orders | Stay at home |
| Close non-essential businesses* |  |
| Suspend non-essential medical services | Suspend non-essential medical services |

^#^ Data on non-pharmaceutical interventions (NPIs) directly targeting transmission reduction

^†^ NPIs groupings used in all analyses; grouped based on correlated presence/absence and semantic similarity.

*Closure of non-essential businesses is grouped with stay at home orders because the effect of closing non-essential businesses is thought to prevent workers from visiting their workplace and most likely lead to them staying home. Closure of non-essential businesses was also highly correlated with stay at home orders (Fig. S11).

**Table S6.** Univariable analysis on county-level characteristics and weekly Rt.

|  | Relative changes in Rt, % (95% CI) | Adjusted R^2^ |
| --- | --- | --- |
| Median household income | -4.3 (-1.4, 10.4) | 0.00% |
| Decile of poverty | 4.7 (3.3, 6.2) | 0.11% |
| Log_10_ population density | 7.1 (5.8, 8.4) | 0.32% |
| Log_10_ population size | 10.6 (9.1, 12.2) | 0.54% |
| Median age | -0.9 (-1.1, -0.7) | 0.27% |
| Decile of percentage of population that is white | -3.9 (-4.4, -3.4) | 0.63% |
| Decile of percentage of population for which highest educational attainment is high school | -1.8 (-2.6, -1.0) | 0.05% |

Table S7. Coefficients estimated in Figure 4.

| **Combination** | ***P*** | **Interventions*** | | | | | | | **Estimates R_eff_ (median, 95% quantiles)** | | | |
| --- | --- | --- | --- | --- | --- | --- | --- | --- | --- | --- | --- | --- |
|  |  | **DC** | **FM** | **LA** | **SH** | **NH** | **MS** | **SC** | | **GEE individual** | **GEE suite** | **XgBoost** |
| 1 | 8.50% | 0 | 0 | 0 | 0 | 0 | 0 | 0 | | 2.70 (1.65, 4.52) | 6.47 (3.91, 9.31) | 4.53 (2.23, 7.26) |
| 2 | 6.50% | 0 | 0 | 0 | 0 | 0 | 0 | 1 | | 1.72 (1.05, 2.87) | 2.08 (1.26, 3.00) | 1.17 (0.55, 5.19) |
| 3 | 4.20% | 0 | 0 | 0 | 0 | 1 | 0 | 1 | | 1.27 (0.77, 2.12) | 1.41 (0.85, 2.03) | 1.04 (0.49, 5.18) |
| 4 | 8.70% | 0 | 1 | 0 | 0 | 1 | 0 | 1 | | 1.04 (0.63, 1.74) | 1.11 (0.67, 1.60) | 1.05 (0.50, 4.91) |
| 5 | 6.30% | 0 | 0 | 1 | 0 | 1 | 0 | 1 | | 1.09 (0.67, 1.83) | 1.14 (0.69, 1.64) | 0.97 (0.44, 4.89) |
| 6 | 4.70% | 0 | 0 | 1 | 1 | 0 | 0 | 1 | | 1.26 (0.77, 2.11) | 1.43 (0.86, 2.06) | 0.91 (0.39, 4.71) |
| 7 | 6.70% | 0 | 0 | 1 | 1 | 1 | 0 | 1 | | 0.93 (0.57, 1.56) | 0.97 (0.58, 1.39) | 0.89 (0.39, 4.72) |
| 8 | 4.30% | 0 | 0 | 1 | 1 | 0 | 1 | 1 | | 1.26 (0.77, 2.11) | 1.42 (0.85, 2.04) | 0.89 (0.37, 4.93) |
| 9 | 4.20% | 0 | 1 | 1 | 0 | 1 | 0 | 1 | | 0.90 (0.55, 1.50) | 0.97 (0.58, 1.39) | 0.97 (0.45, 5.01) |
| 10 | 4.50% | 1 | 0 | 1 | 1 | 1 | 1 | 1 | | 0.65 (0.39, 1.08) | 0.78 (0.47, 1.13) | 0.85 (0.36, 4.95) |

* DC, daycare closure; FM, face mask; LA, leisure activities closure; SH, stay home; NH, nursing home visit ban; MS, medical service suspension; SC, school closure,

### Supplementary Figures

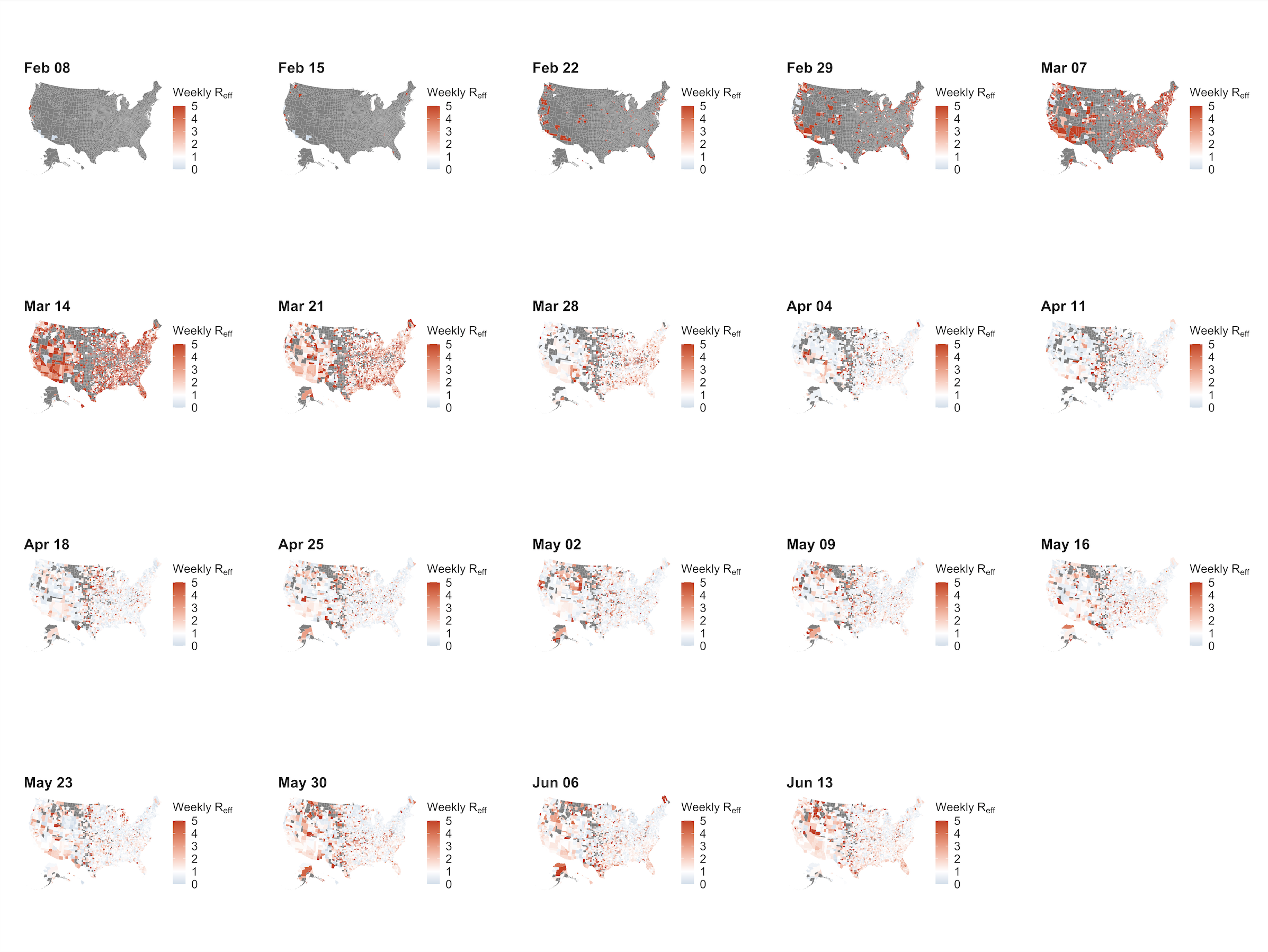

**Figure S1. Maps of weekly reproduction number.** Grey indicates no data available.

**Figure S2. Temporal distribution of intervention by state.** Shown for weeks when R_eff_ estimates were available.
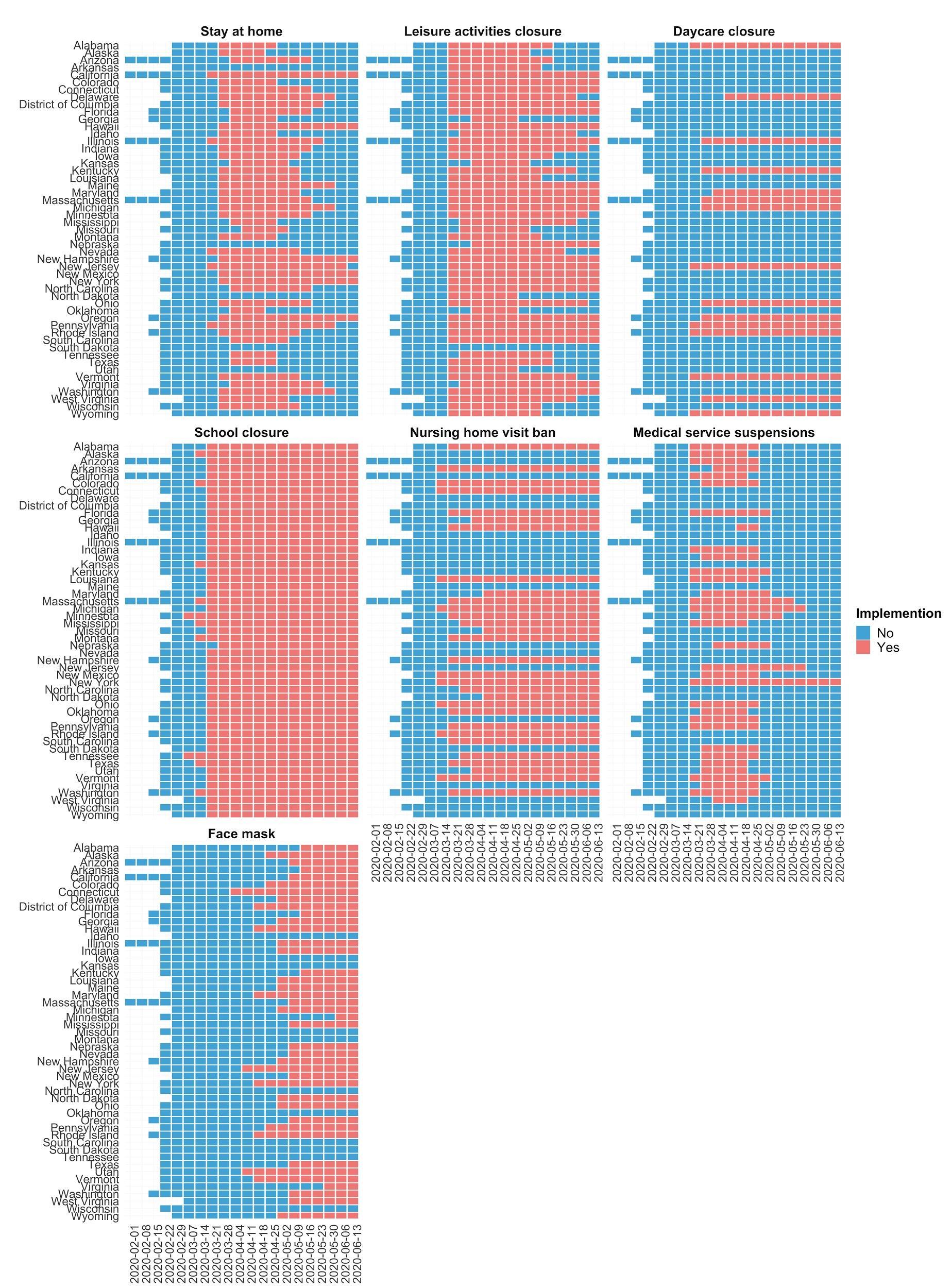

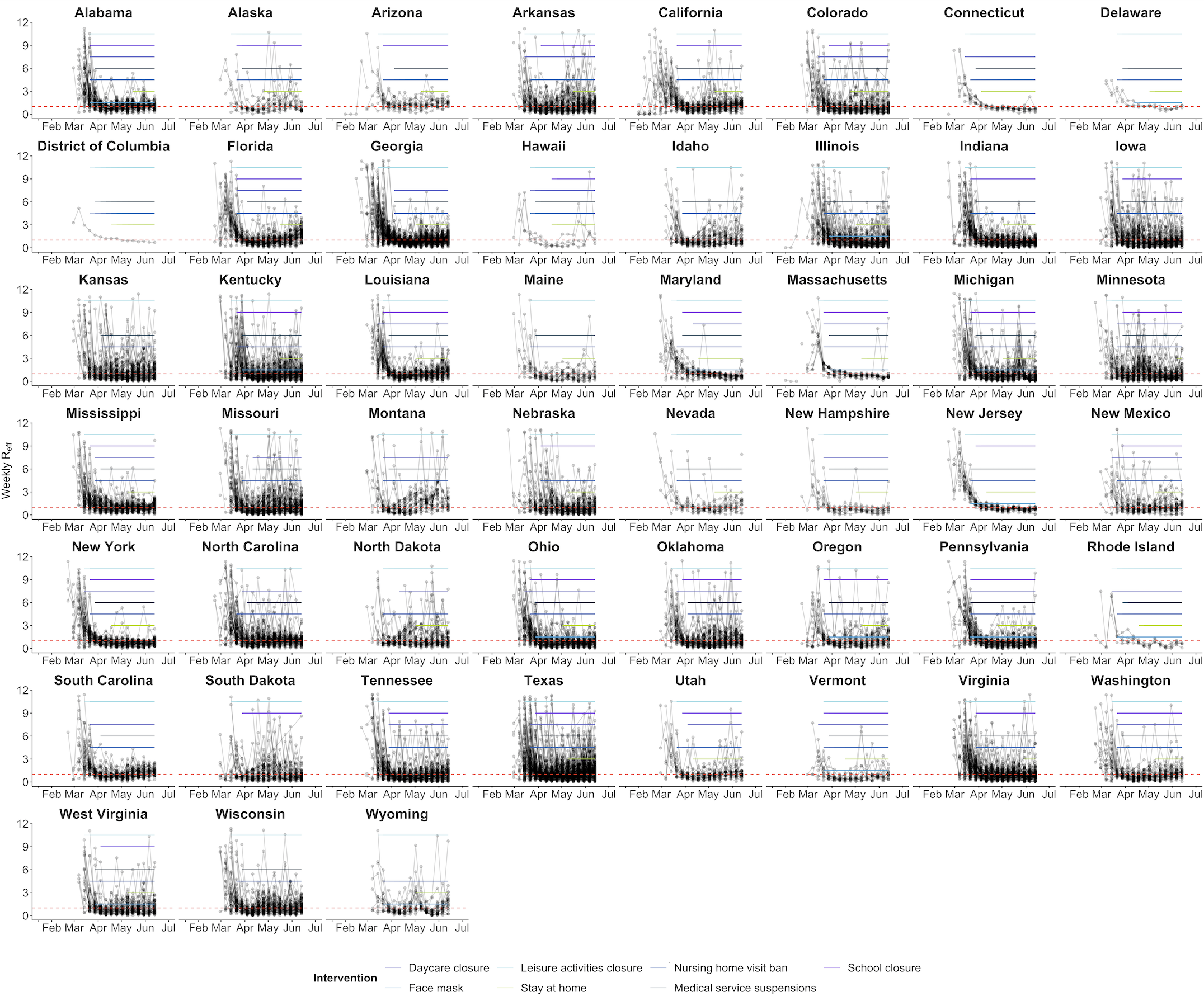

**Figure S3: Estimates of R_eff_ estimates from confirmed cases from each county organized by state.** County-specific time series of R_eff_ are shown. Colored solid lines indicate the duration of interventions implementation in each state. Red dashed line indicates a reproductive number of one.

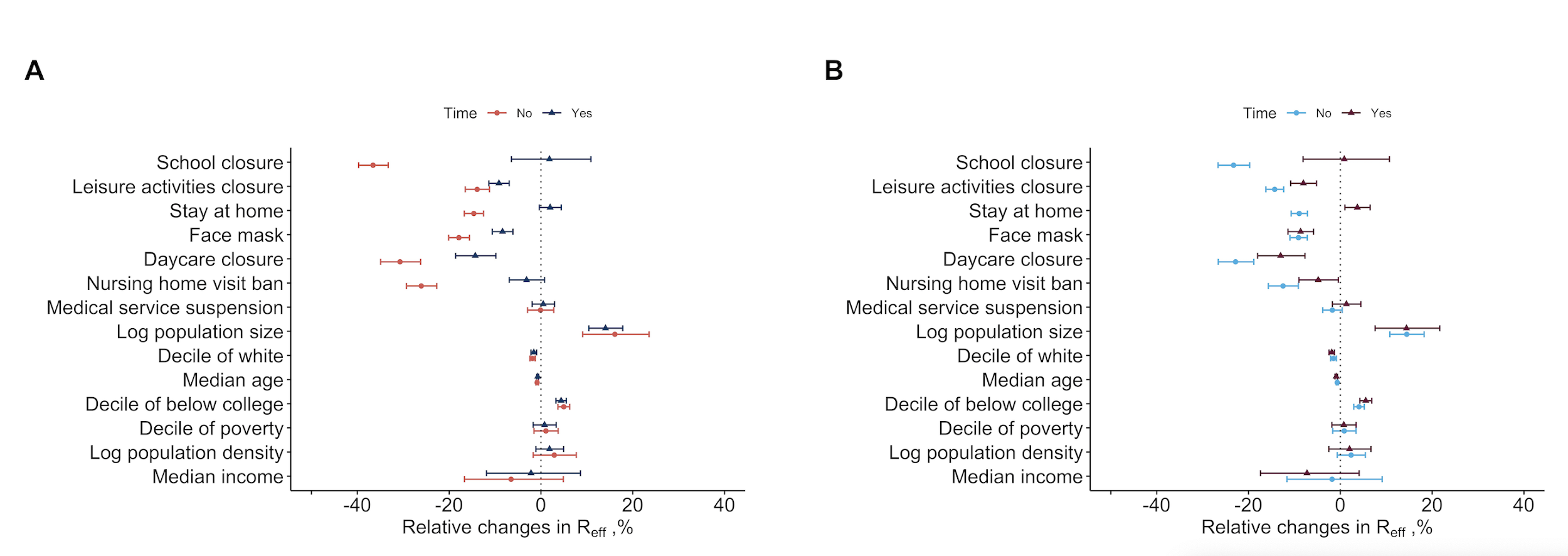

**Figure S4. Effects of non-pharmaceutical interventions (NPIs) estimated from generalized estimating equations (GEEs) (A) and lag model regression (B) with and without calendar time.** Calendar time was modelled as categorical variables, which indicate the number of weeks since the United States saw its first case. The shown models without calendar time are main models fitted with GEEs (A) and lag model regression (B) (Table S2).

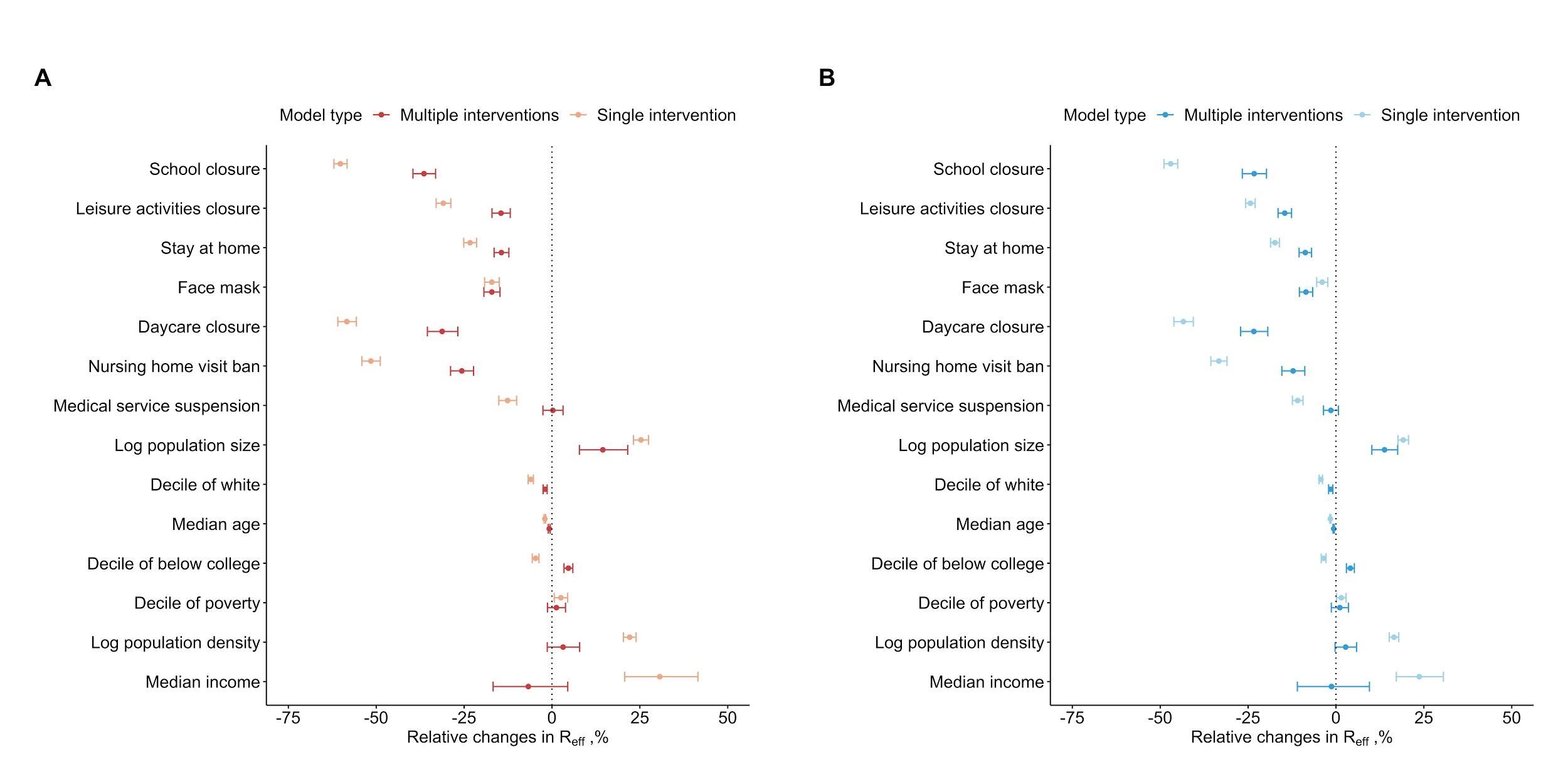

**Figure S5. Comparison of effects of non-pharmaceutical interventions (NPIs) on transmission that were estimated from single-intervention (univariate) and multi-intervention models (multivariate) (main models).** For single-intervention models, we added one intervention to the base model (Table S2) at a time. Models were fitted with GEEs (A) and lag model regression (B).

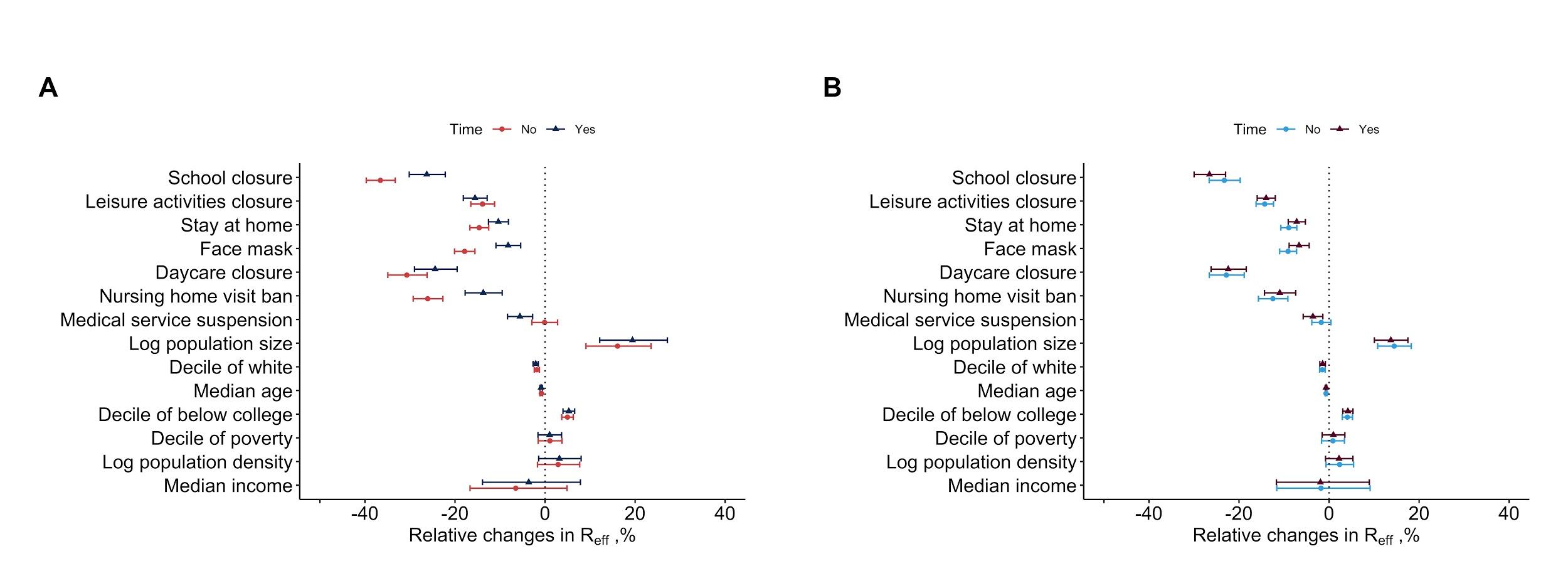

**Figure S6. Effects of non-pharmaceutical interventions (NPIs) estimated from generalized estimating equations (GEEs) (A) and lag model regression (B) with and without county-specific time.** County-specific time was modelled as categorical variables, which indicate the number of weeks since the county saw its first case (Table S2; see Methods). The shown models without county-specific time are main models fitted with GEEs (A) and lag model regression (B) (Table S2).

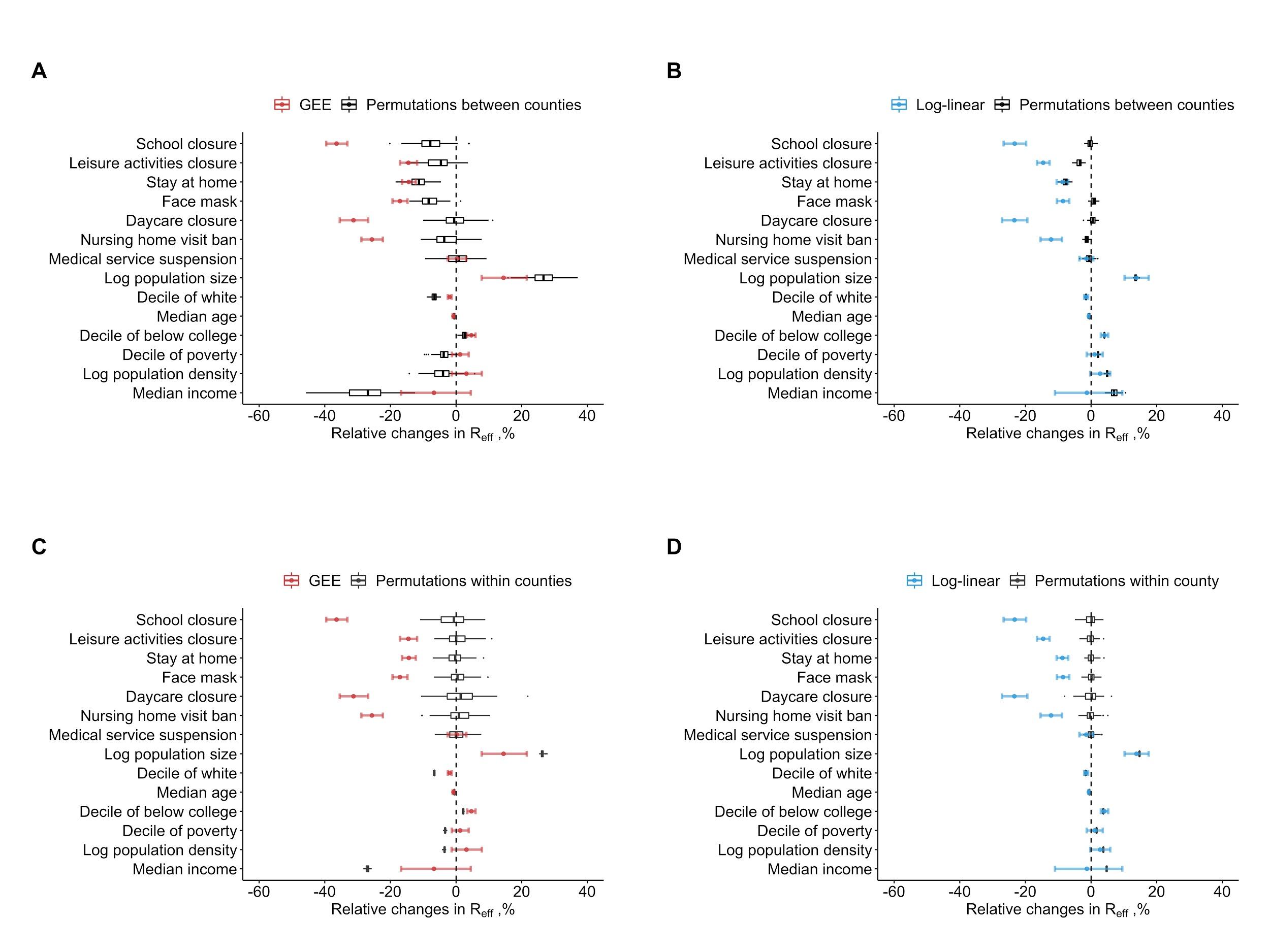

**Figure S7. Effects of non-pharmaceutical interventions (NPIs) estimated from the main models compared against estimates when county-level NPIs suites were permuted spatially (A, B for GEE and lag model, respectively) or temporally (C, D for GEE and lag model regression respectively).**

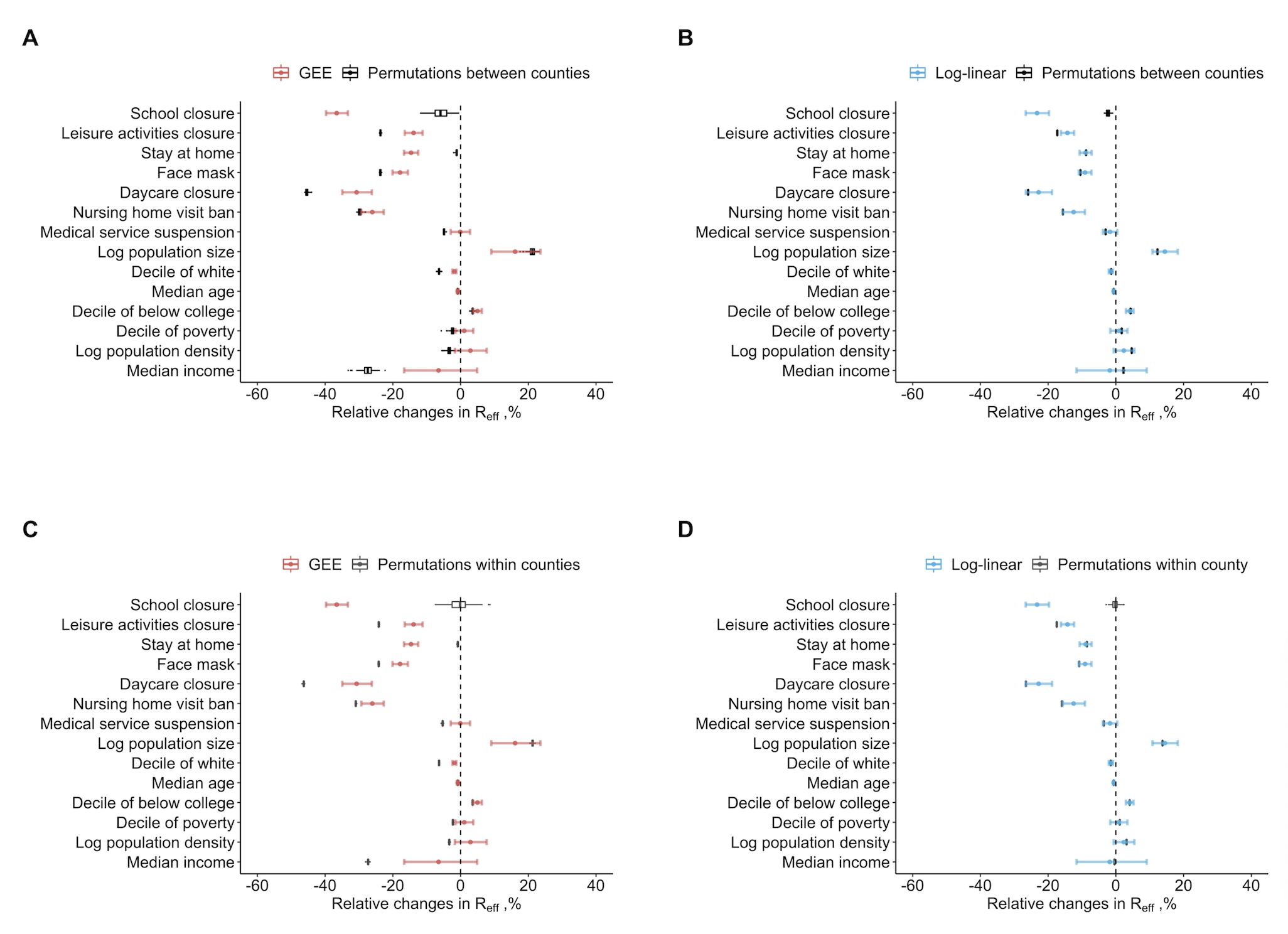

**Figure S7 Alt. Effects of non-pharmaceutical interventions (NPIs) estimated from the main models compared against estimates when county-level school closures were permuted spatially (A, B for GEE and lag model, respectively) or temporally (C, D for GEE and lag model regression respectively).**

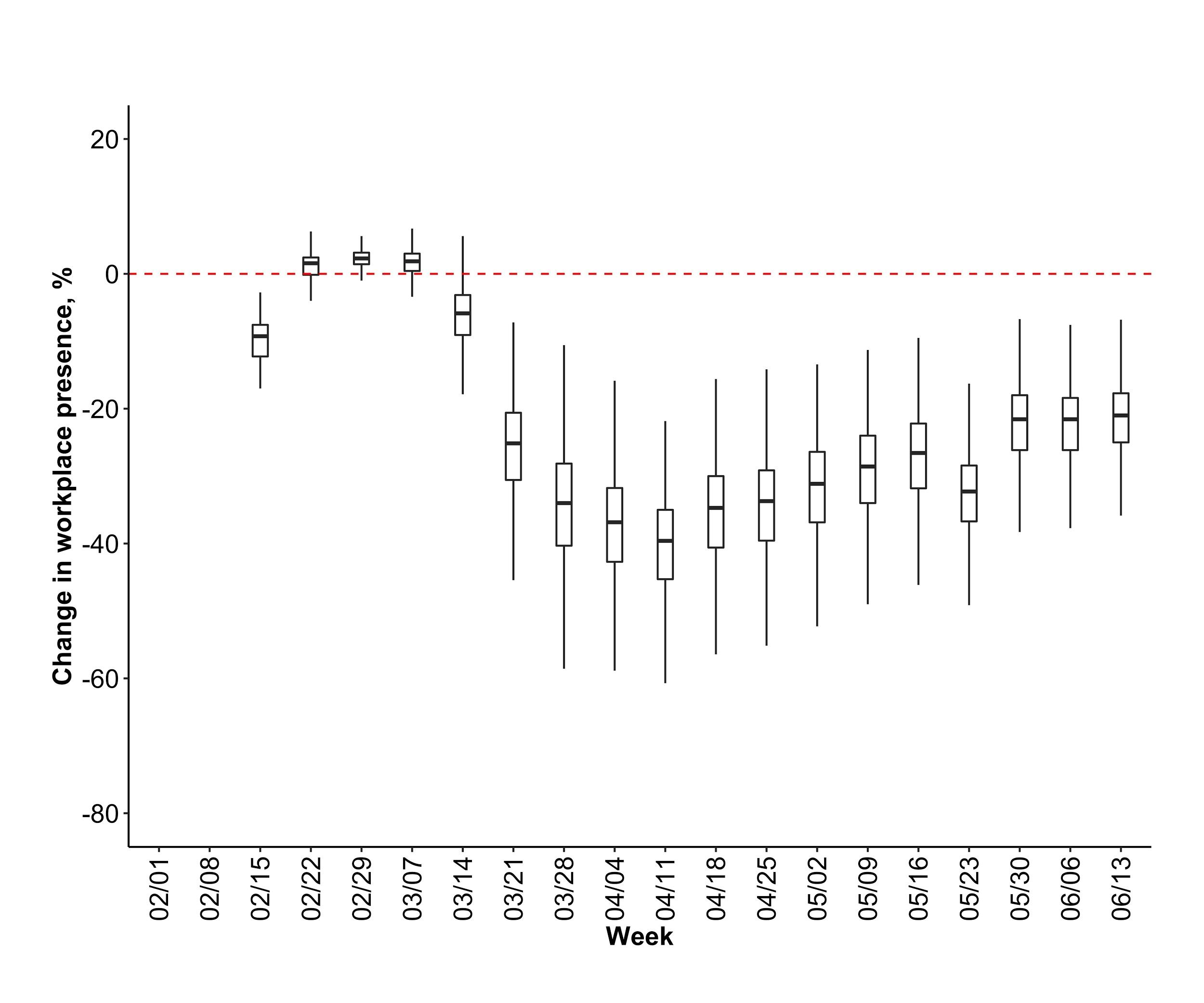

**Figure S8. Temporal distribution of changes in workplace presence from Google data.** Data on workplace presence relative to pre-pandemic periods (*6*).

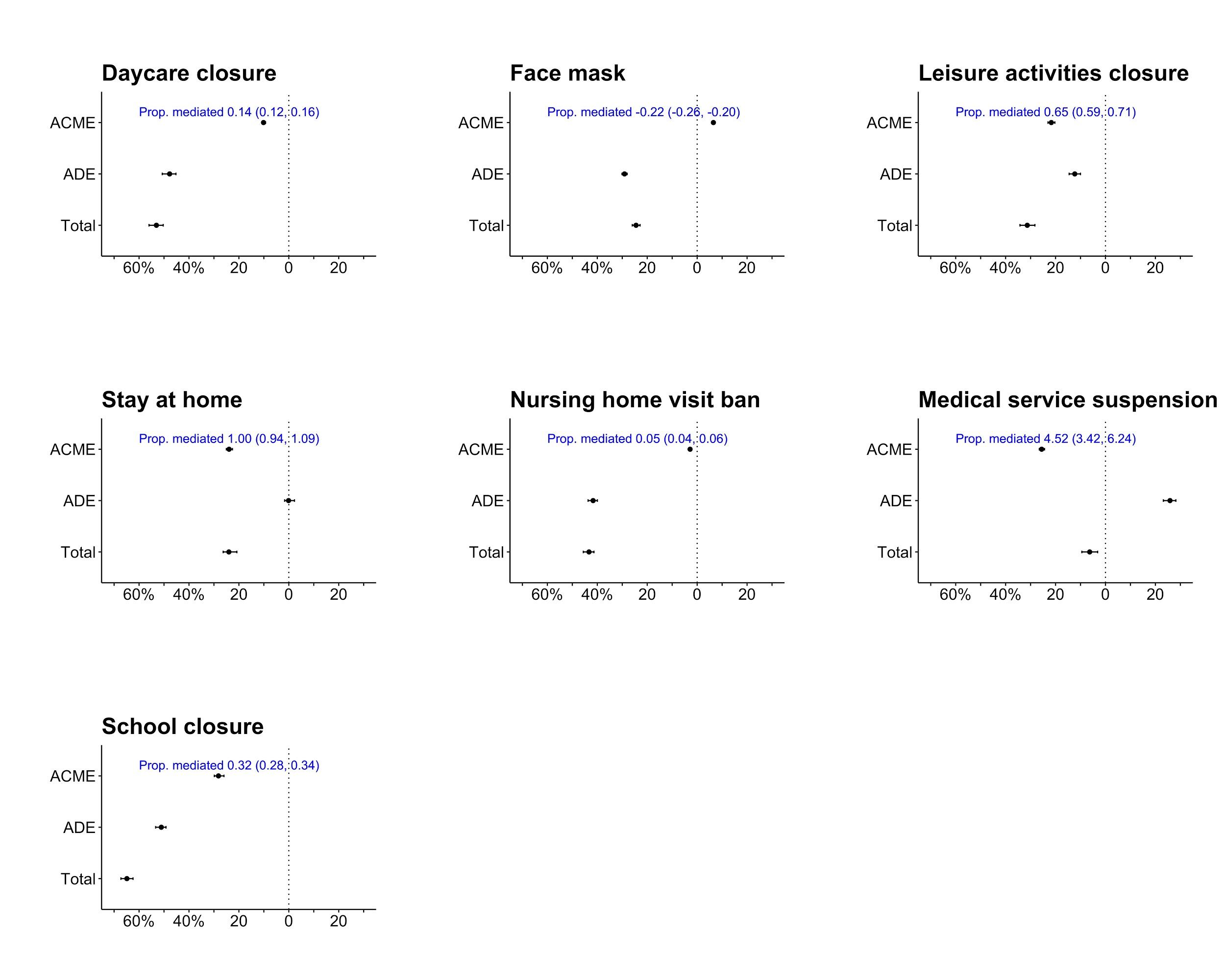
**Figure S9. Mediation analyses of workplace presence on the association between non-pharmaceutical interventions (NPIs) and SARS-CoV-2 transmissions.** Models were fitted in log-linear regression and adjusted for county-level characteristics and autocorrelation of log R_eff_. ACME, average causal mediation effect; ADE, average direct effect; Total, total effect.

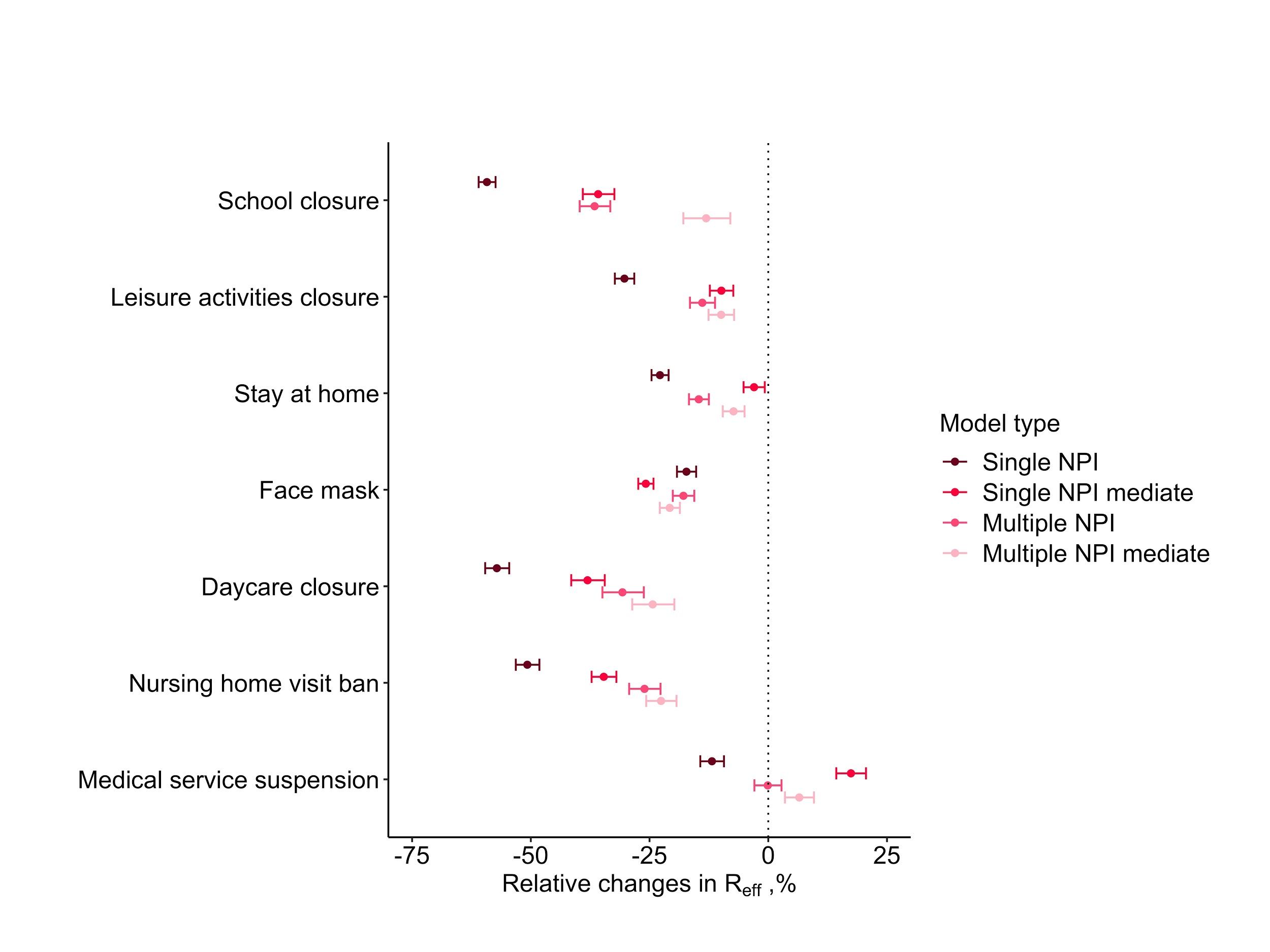

**Figure S10. Effects of workplace presence on the association between non-pharmaceutical interventions (NPIs) and SARS-CoV-2 transmissions.** All models were fitted with GEEs and adjusted for county-level characteristics and autocorrelation of log R_eff._ Single NPI model includes the examined NPI; single NPI mediate model includes the examined NPI and the workplace presence; multiple NPI mediate model includes all NPIs and the workplace presence.

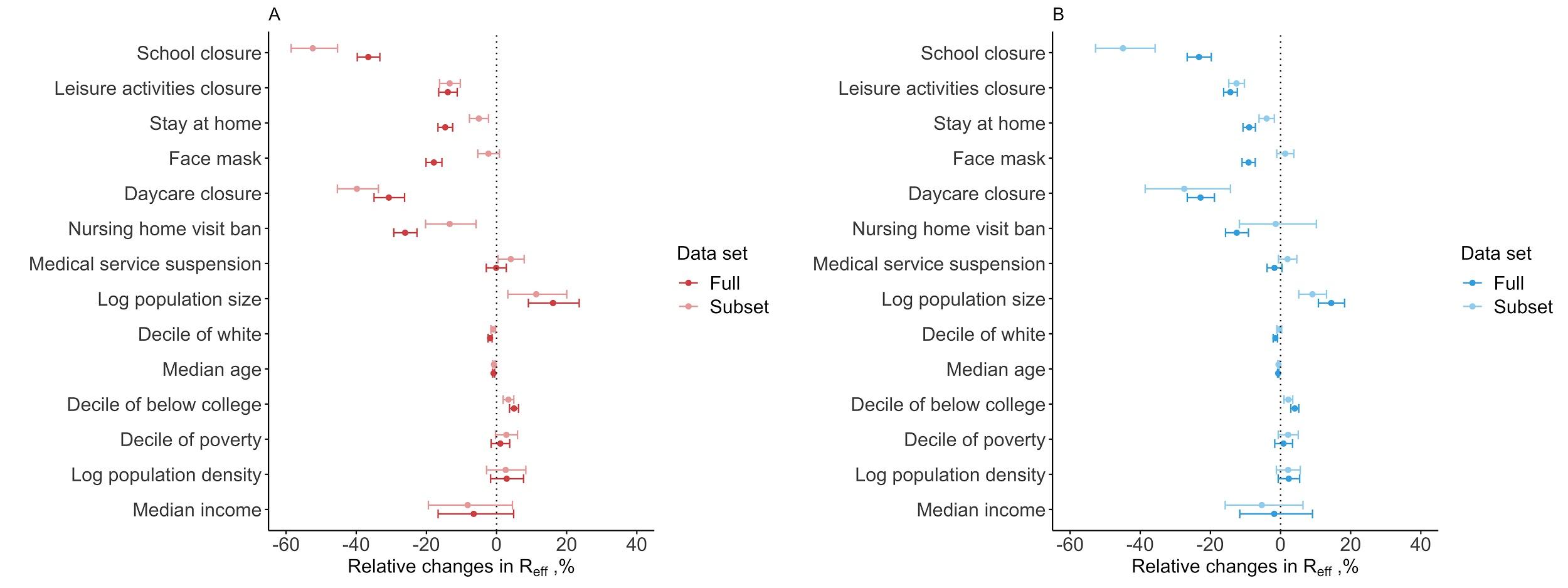

**Figure S11. Effects of non-pharmaceutical interventions (NPIs) estimated with R_eff_ estimates subsetted to two weeks after the county's first case onwards compared to the full dataset from generalized estimating equations (A) and lag model regression (B).**

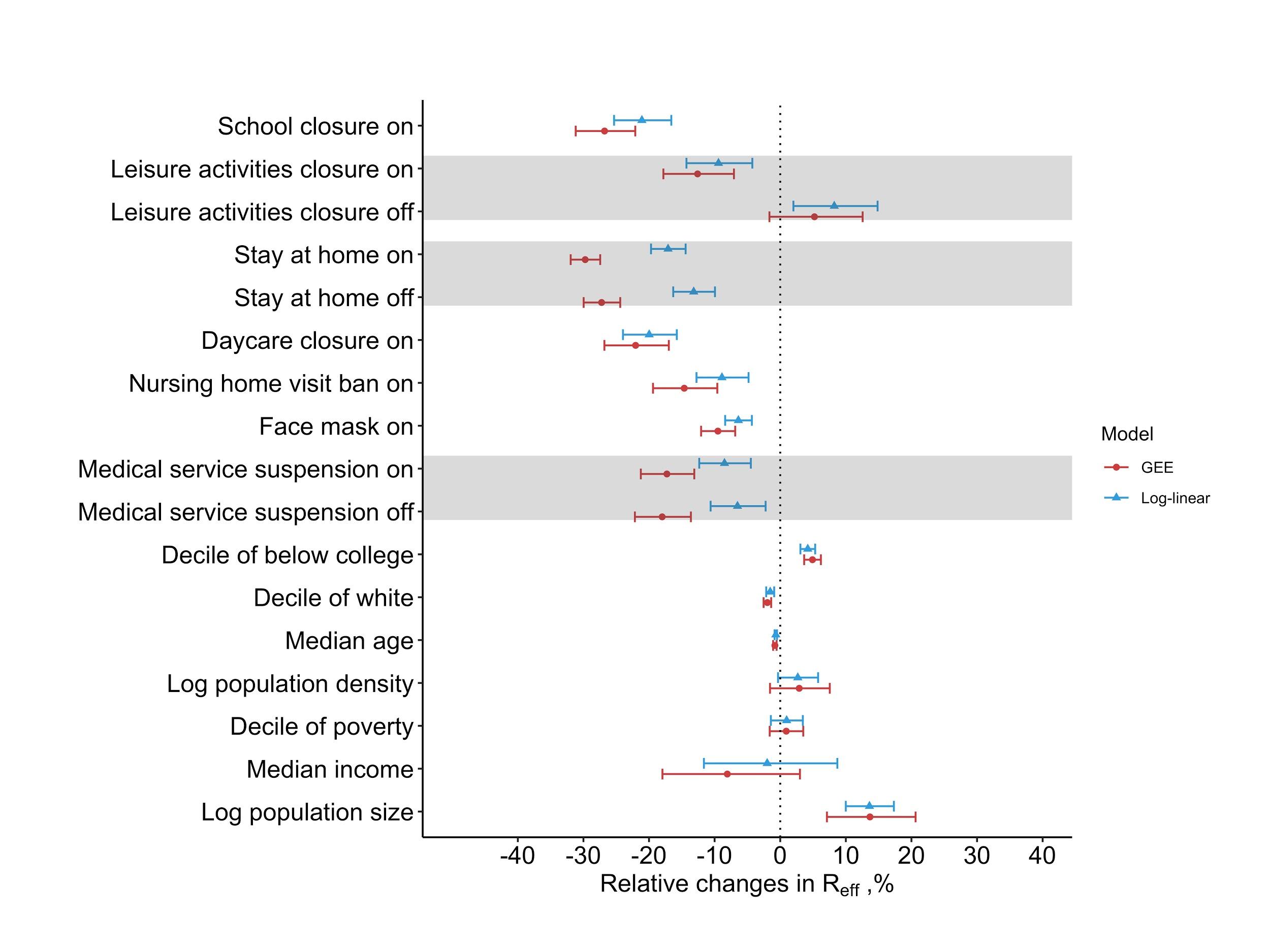

### Figure S12. Effects of relaxing non-pharmaceutical interventions (NPIs) on transmission. Main models were fitted by adding additional covariates to indicate the relaxation of NPIs when applicable (i.e. leisure activities closure, stay at home order and medical service suspension).

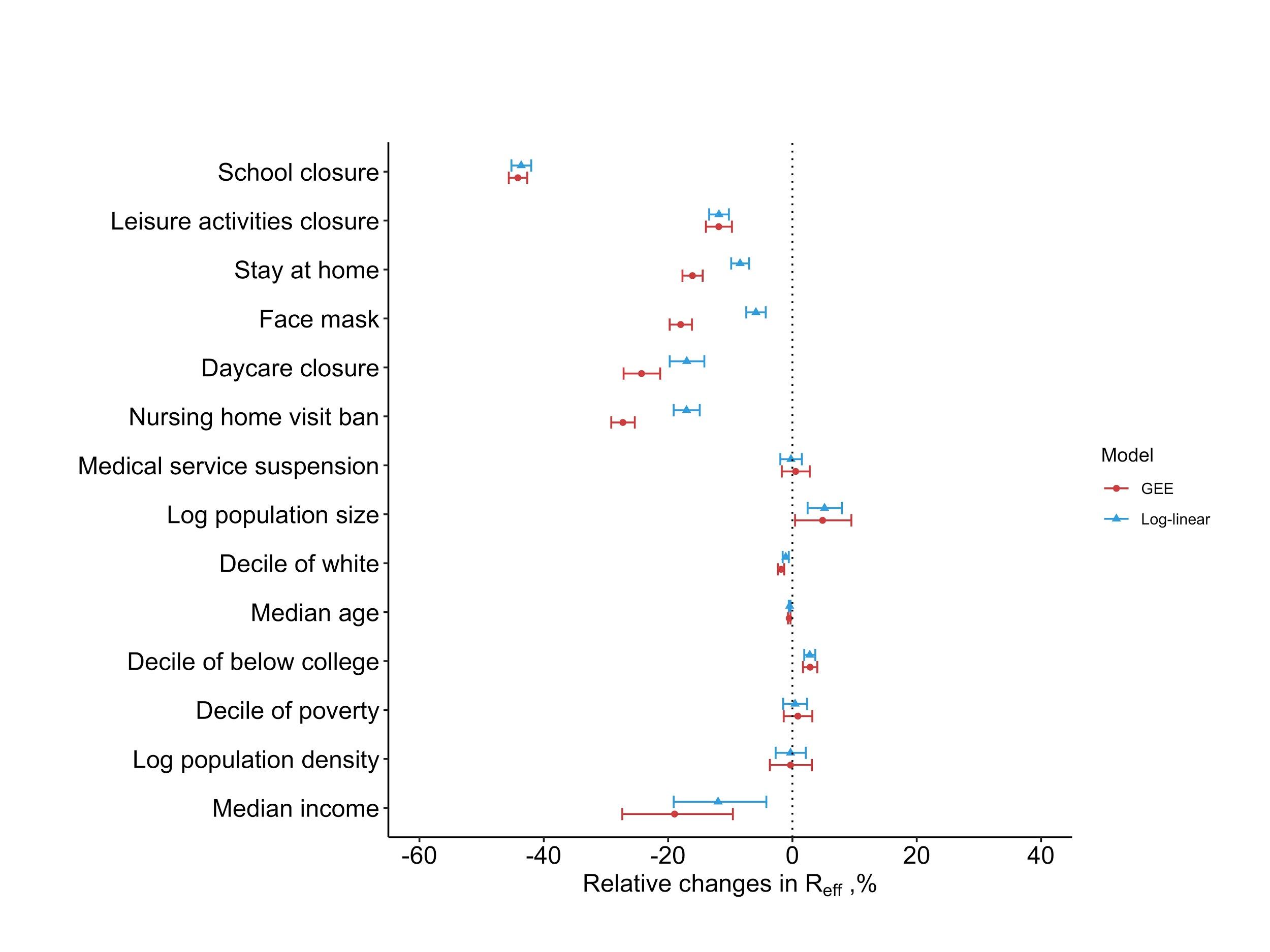

### Figure S13. Effects of non-pharmaceutical interventions (NPIs) on transmission, using reproduction numbers (R_eff_) that were estimated from stochastic reconstruction of infections from cases by sampling delay distribution of time from infection to confirmation (as opposed to deconvolution method used in main text).

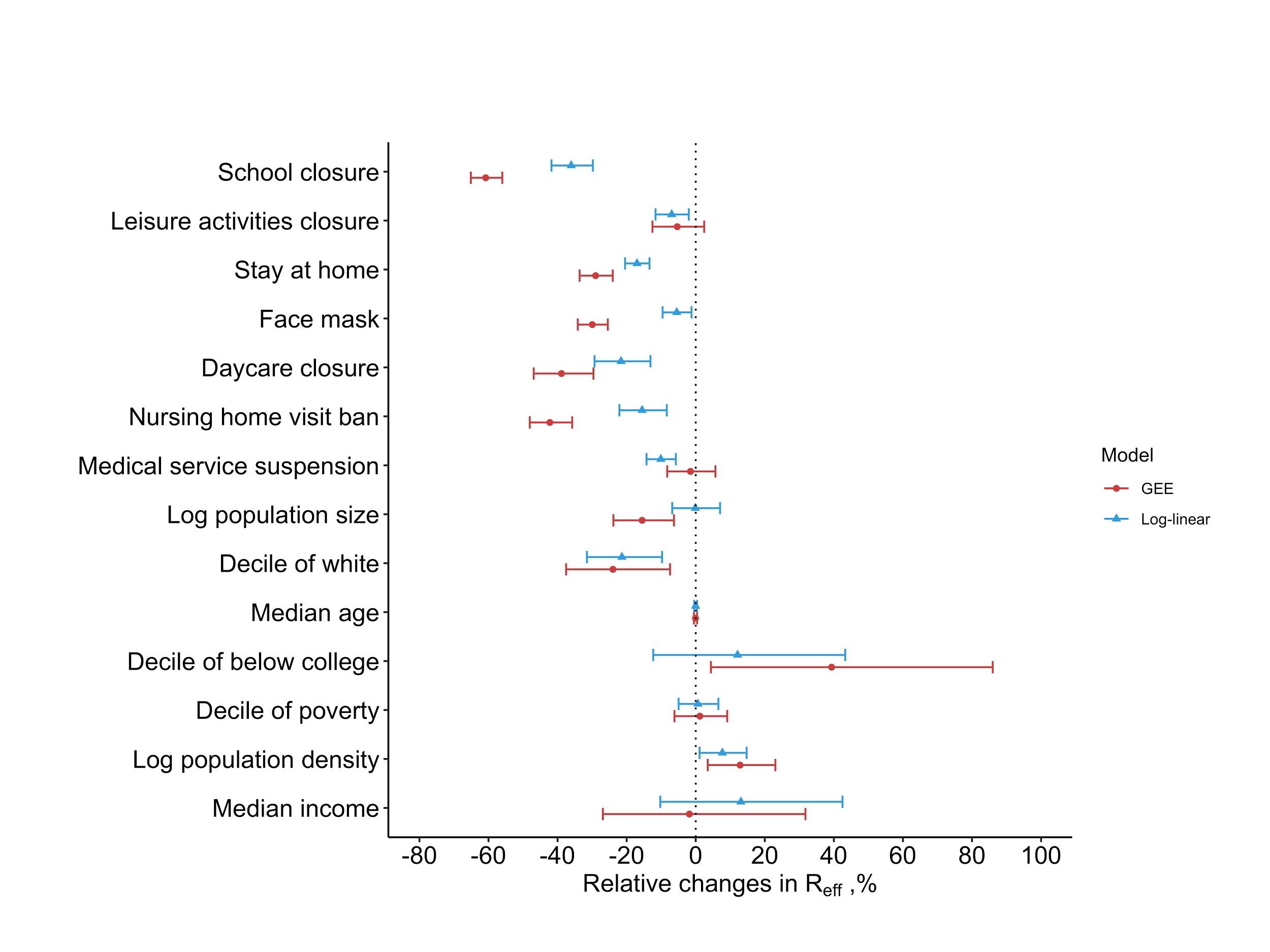

**Figure S14. Effects of non-pharmaceutical interventions (NPIs) on transmission, using reproduction numbers (R_eff_) estimated from deconvolution of county-level COVID-19 death reports.**

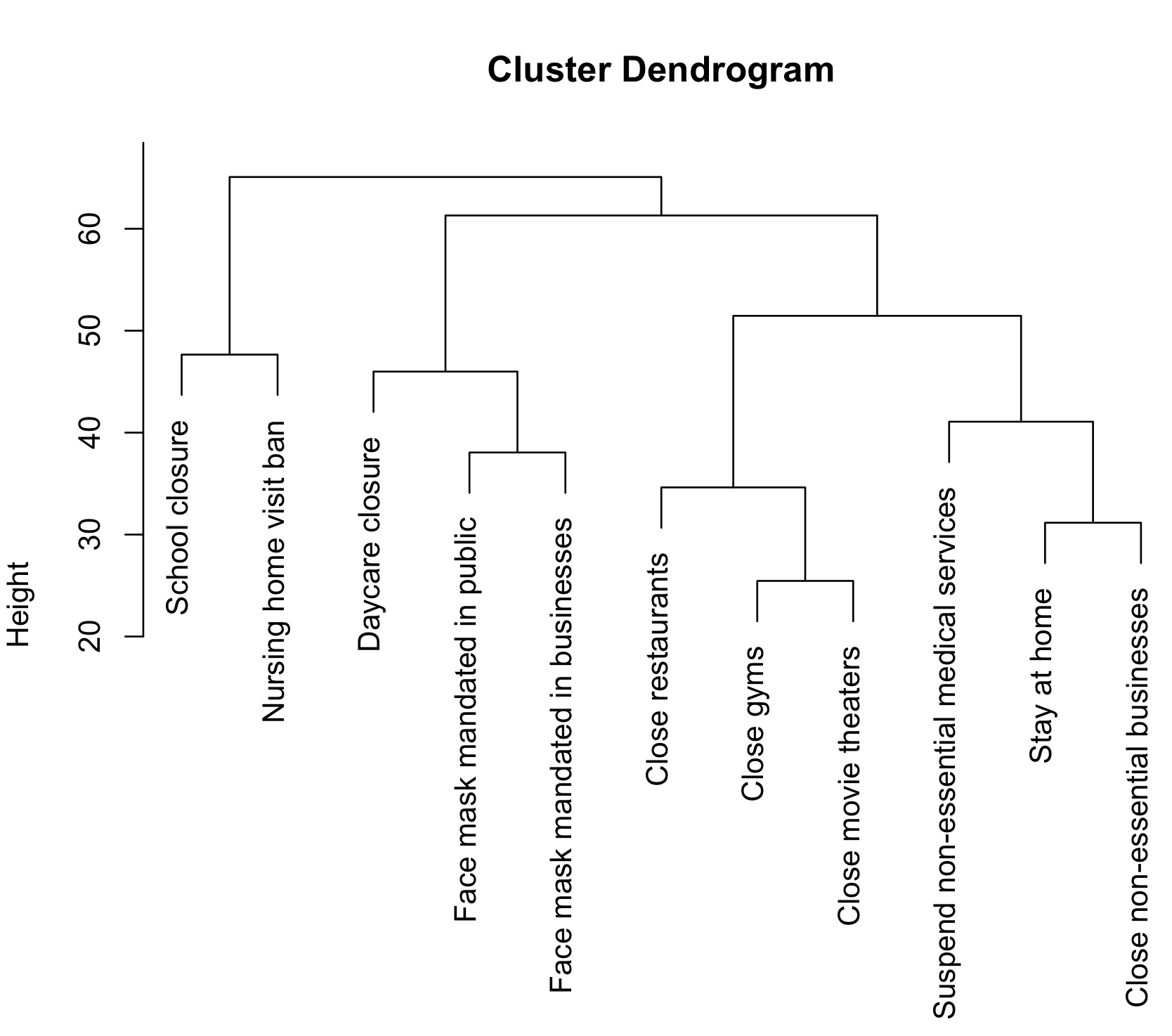

**Figure S15. Hierarchical clustering of non-pharmaceutical interventions (NPIs) occurrences in time.**
